# *qbaconfound*: A flexible Monte Carlo probabilistic bias analysis for unmeasured confounding

**DOI:** 10.1101/2025.08.12.25333217

**Authors:** Emily Kawabata, Chin Yang Shapland, Tom Palmer, David Carslake, Kate Tilling, Rachael Hughes

## Abstract

**Background:** Unmeasured confounding is a persistent concern in observational studies. We can quantitatively assess the impact of unmeasured confounding using a quantitative bias analysis (QBA). A QBA specifies the relationship between the unmeasured confounder(s), *U*, and study data via its bias parameters.

There are two broad classes of QBA methods: deterministic and probabilistic. We focus on a probabilistic QBA which incorporates external information about *U* via prior distribution(s) placed on these bias parameters and can be implemented as a Bayesian QBA or a Monte Carlo QBA. A Bayesian QBA combines the prior distribution(s) with the data’s likelihood function whilst a Monte Carlo QBA samples the bias parameters directly from their prior distributions. Software implementations of probabilistic QBAs to unmeasured confounding are scarce and mainly limited to unadjusted analyses of a binary exposure and outcome. One exception is R package *unmconf* (Hebdon et al 2024, BMC Med. Res. Methodol., https://doi.org/10.1186/s12874-024-02322-2) which implements a Bayesian QBA, applicable when the analysis is a generalised linear model (GLM). However, for a study with *q* measured confounders and a single *U, unmconf* requires information on at least 3+ *q* bias parameters, which is burdensome when *q* >1 and validation data are unavailable.

**Aim:** We propose a flexible Monte Carlo QBA where the number of bias parameters is independent of the number of measured confounders. It is applicable to a GLM or survival proportional hazards model, with binary, continuous, or categorical exposure and measured confounders, and one or multiple (≥ 2) binary or continuous unmeasured confounders.

**Methods:** Via simulations, we evaluated our Monte Carlo QBA for different analyses (e.g., varying the regression model, type of variables for the exposure and unmeasured confounder), and different levels of dependency between the measured and unmeasured confounders. Also, using our proposed bias model, we compare a Monte Carlo implementation to a fully Bayesian implementation when the analysis is a linear or logistic regression. We repeat the simulation study for prior distributions with different levels of informativeness.

**Results:** Ignoring *U* resulted in substantially biased estimates with substantial confidence interval undercoverage (e.g., 57%). Our Monte Carlo QBA (with informative priors) resulted in unbiased (or minimally biased) point estimates and interval estimates with close to nominal coverage. For binary *U*, levels of bias were marginally higher when *U* was strongly correlated with the measured confounders. The performances of the Monte Carlo and Bayesian implementations were comparable.

**Conclusion:** We have proposed a flexible probabilistic QBA for unmeasured confounding which is applicable for a wide range of regression-based analyses. We have minimised the burden placed on the user by limiting the number of bias parameters and avoiding the need for specialist knowledge about Bayesian inference or Bayesian software. Our proposed Monte Carlo QBA will be implemented as Stata command and R package, *qbaconfound*.

## 1. Introduction

The main aim of many epidemiology studies is to estimate the causal effect of an exposure (or a treatment) on an outcome (here onward, shortened to *exposure effect*). Unmeasured confounding is a common problem in observational studies because participants are not randomised to exposure (or treatment) groups. Commonly used statistical methods, such as standardization, inverse probability weighting, regression adjustment, g-estimation, stratification and matching, assume “no unmeasured confounding” (i.e., the adjustment model is correct, and a sufficient set of confounders has been measured without error [1]). Failure to appropriately account for unmeasured or poorly measured confounders in analyses may lead to invalid inference [2, 3, 4].

There are several approaches to assess causality which depend on assumptions other than “no unmeasured confounding” (e.g., self-controlled study designs, prior event rate ratio, instrumental variable analysis, negative controls, and perturbation variable analysis, and methods that use confounder data collected on a study sub-sample [5]). When none of these approaches are applicable (e.g., study lacks an appropriate instrument or subsample data on the unmeasured confounders) then the analyst should assess the sensitivity of the study’s conclusions to the assumption of no unmeasured confounding using a quantitative bias analysis (QBA). Note that a QBA may be conducted to quantify the potential impact of other sources of bias such as nonignorable missing data, information bias, and selection bias (e.g., [6, 7, 8]). From here onwards, we shall use the term *QBA* to refer to a QBA for unmeasured confounding.

A QBA can be used to quantify the likely magnitude and direction of the bias under different plausible assumptions about the unmeasured confounder(s), usually assuming no other sources of bias. Generally, a QBA requires a model (known as a bias model) for the observed data and the unmeasured confounders and will include one or more parameters (known as bias or sensitivity parameters) which cannot be estimated from the observed data. Therefore, values for these bias parameters must be prespecified before conducting the QBA. These bias parameter values are obtained from external sources such as external validation studies, published literature, or expert opinion [9].

There are two broad classes of QBA methods: deterministic and probabilistic [10]. We focus on a probabilistic QBA which specifies a prior probability distribution for the bias parameters to explicitly model the analyst’s assumptions about which combinations of the bias parameters are most likely to occur and to incorporate their uncertainty about their true values [10, 11]. A probabilistic QBA can be implemented as a fully Bayesian analysis (where the prior distribution is combined with the observed data’s likelihood function) or using the Monte Carlo approach (which repeatedly samples the bias parameters directly from their prior distributions then fixes the bias parameters to these drawn values to estimate the exposure effect) [11, 6]. Both types of probabilistic QBA generate a distribution of bias-adjusted estimates which is then summarised to give a point estimate (i.e., the most likely bias-adjusted estimate under the QBA’s assumptions) and an interval estimate (i.e., defined to contain the true exposure effect with a prespecified probability) which accounts for uncertainty due to the unmeasured confounding and sampling variability [12].

Currently, QBA methods are not routinely used in practice [13]. A study published in 2024 concluded that the “application of QBA is rare in the literature but is increasing over time” [14]. A lack of knowledge about QBA, and of analyst-friendly methods and software have been identified as barriers to the widespread implementation of a QBA [12, 9, 15]. A review of software implementations of QBA for unmeasured confounding reported that most programs were restricted to analyses with a binary or continuous outcome and exposure and identified a need for software implementations of probabilistic QBAs and in platforms other than R [16].

Software implementations of a probabilistic QBA that were not reported by the review [16] (e.g., due to their release outside the review window) include R packages *unmconf* [17], *CIMTx* [18], *SAMTx* [19], and *episensr* [20], shiny application *apisensr* [21], and Stata command *episens* [22]. Programs *episensr, apisensr*, and *episens* all implement a Monte Carlo QBA using the same bias model (proposed by Fox et al [12]) which is restricted to the simple setting of 2 × 2 tabulations (i.e., a binary exposure, binary outcome and no measured confounders). Programs *CIMTx* and *SAMTx* also implement a Monte Carlo QBA but are applicable only when the outcome is binary, and the exposure is categorical (measured confounders are allowed). The most recent addition is R package *unmconf* which implements a Bayesian bias analysis and is applicable to a wider range of analyses (specifically, different types of generalised linear models (GLMs)). Also, unlike most software implementations, the bias model of *unmconf* does not assume independence between the unmeasured and measured confounders and allows for two correlated unmeasured confounders (as opposed to the conservative approach of modelling a linear combination of “independent” unmeasured confounders). A notable feature of *unmconf* is that the number of bias parameters depends on the number of measured confounders; for example, a logistic regression analysis with two binary unmeasured confounders and 12 continuous measured confounders requires an additional 20 bias parameters compared to the same analysis with only 2 continuous measured confounders (i.e., 31 bias parameters for the former versus 11 for the latter). Defining informative priors for 10 or more bias parameters places a huge burden on the user and may reflect why the realistic examples (i.e., allowing for measured confounders) illustrating *unmconf* are in situations where validation data are available, and so vague priors can be specified for all or a large majority of the bias parameters [23, 24].

We propose a Monte Carlo QBA that retains the flexibility of *unmconf* but uses far fewer bias parameters. Our proposed bias model is applicable to a GLM or survival proportional hazards model and allows for: (i) a binary, continuous, or categorical exposure and measured confounders, (ii) correlation between the measured and unmeasured confounders, and (iii) one or multiple (≥ 2) binary or continuous unmeasured confounders. In section 2, we describe the bias model of *unmconf* followed by a description of our proposed bias model and Monte Carlo implementation. Section 3 describes the simulation study where we evaluate our proposed Monte Carlo QBA for different types of analyses, and different levels of dependency between the measured and unmeasured confounders. Also, via simulations and a real data analysis we compare a Monte Carlo implementation of our proposed bias model to a fully Bayesian analysis (sections 3 and 4, respectively). We conclude with a discussion in section 5.

## 2. Probabilistic bias analysis for confounding

### 2.1 Set-up

We want to estimate the causal effect of an exposure on an outcome. Without any loss of generality, we shall focus on a binary or continuous exposure and direct the reader to supplementary section 4 for the scenario with a categorical exposure. For subject *i* (*i*= 1, …, *n*), let *Y*_*i*_ denote the outcome and *X*_*i*_ denote a binary or continuous exposure. The exposure-outcome association is confounded by measured and unmeasured confounders. For *q*^*^ measured confounders and *q* ≥ *q*^*^, let *C*_*i*_ = *C*_*i*,1_, ‥, *c*_*i,q*_) denote a *q*-vector of corresponding variables, where *c*_*i,j*_ may be continuous (linear, polynomial or fractional polynomial variable of a continuous measured confounder), a binary measured confounder, a dummy variable of a categorical measured confounder, or an interaction term between two measured confounders (*j* = 1, …, *q*). Also, let *U*_i_ = (*U*_*i*,1_, ‥, *U*_*i,p*_) denote a *p*-vector of binary or continuous unmeasured confounders (included as linear terms only). We assume that, for *i*= 1,…, *n*, (*Y*_*i*_, *X*_*i*_, *C*_*i*_, *U*_*i*_) are independent and identically distributed draws from a joint probability distribution. For ease of reading, we have dropped the notation for subject *i* from here onwards.

The substantive analysis of interest, *Y*|*X, C, U*, is a regression of *Y* on *X* adjusted for *C* and *U*.

It may be a generalised linear model (GLM), 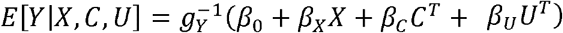 with link function 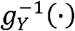, or a Cox proportional hazards model, *h*(*S*|*X, C, U* = *h*_*0*_ (*S*) *exp*(*β* _*X*_*X*+ *β* _C_*C*^*T*^ + *β* _*U*_*U*^*T*^ with hazard function and baseline hazard *h*(·) and *h*_0_ (·), respectively, and *Y*= (*S, D*) for survival time *S* and event indicator *D* (1 for events, 0 for censored observations). The exposure effect of interest is *β* _*X*_ and vectors 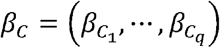 and 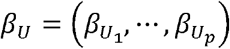 denote the coefficients corresponding to the measured and unmeasured confounders, respectively. We assume the substantive analysis is correctly specified and would give unbiased results if *U* was measured.

We shall refer to *Y*|*X, C, U* as the *full model* and its estimate of *β* _*X*_ as the *full model* estimate. The *naïve model* is the same type of regression as the substantive analysis but with *U* omitted (i.e., a regression of *Y* on *X* adjusted for *C* only) and we refer to its estimate of *β* _*X*_ as the *naïve model* estimate. See supplementary table 1 for a summary of the notation.

**Table 1:**
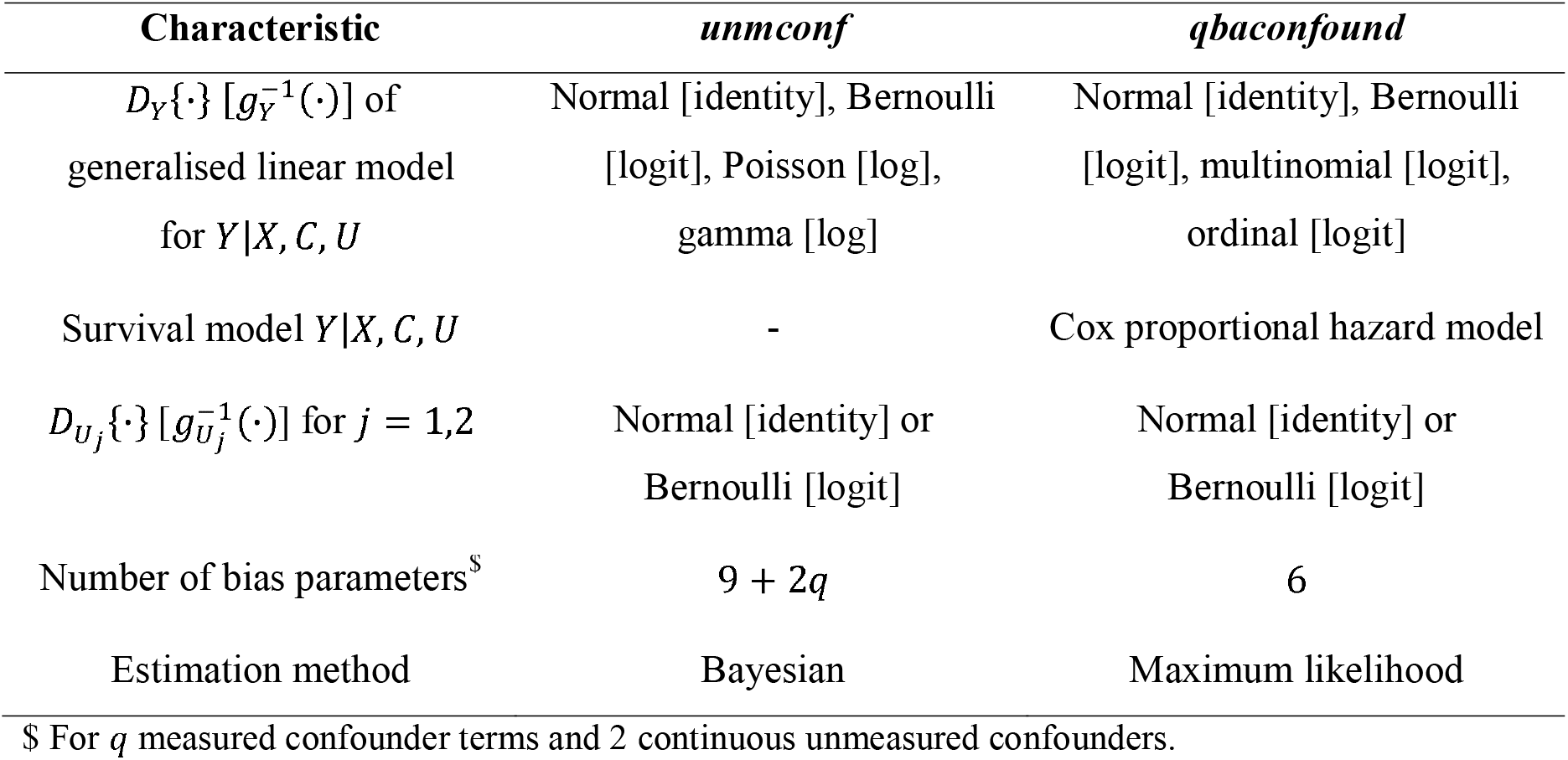
Summary of the main features of *unmconf* and *qbaconfound*

### 2.2 Bayesian QBA unmconf

Hebdon et al [23] propose a Bayesian QBA implemented as R package *unmconf* [17], which is applicable when the substantive analysis is a linear, logistic, Poisson, or Gamma regression (i.e., *Y*∼*Gamma*{·} with a log link) and there are one or multiple (*m*_*U*_ ≥2) continuous or binary unmeasured confounders. Following Hebdon et al and the specifications of the R package, we describe the bias model of *unmconf* with respect to *m*_*U*_ =2 bias parameters. [Note that a user can modify the JAGS code generated by *unmconf* to allow for more than two unmeasured confounders]. The bias model of *unmconf* specifies the conditional joint distribution of *Y* and *U*= (*U*_1_, *U*_2_) given *X* and *C* as:

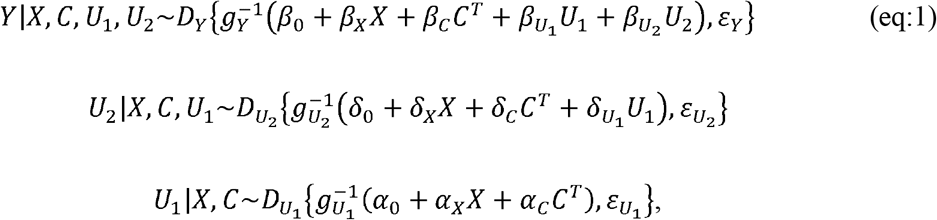

where *U*_2_|*X, C, U*_1_ and *U*_1_|*X, C* are linear or logistic regressions. For these GLMs, 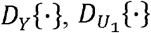, and 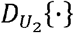 denote response distributions of Y, *U*_1_ and *U*_2_, respectively, and 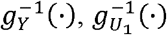, and 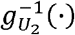 denote the corresponding link functions. When there is a single unmeasured confounder, regression *U*_2_|*X, C, U*_1_ is omitted. Parameters 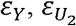, and 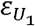 are not part of the linear predictor and are only present for certain GLMs (such as the residual variance in a linear regression). The bias parameters are coefficients 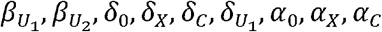 and in some cases additional parameters 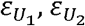 Therefore, for *q* measured confounder terms, the minimum number of bias parameters is 3+ *q* for a single unmeasured confounder and 7+ 2*q* for two unmeasured confounders.

The *unmconf* bias model, eq:1, is estimated within the Bayesian framework. When validation data are available vague prior distributions can be specified for all parameters (including the bias parameters) such as *N* (0,1000) for coefficients of a linear regression and *Gamma* (0.001, 0.001) (i.e., shape and rate of 0.001) for the inverse of the residual variance. In the absence of validation data, informative prior distributions are required for the bias parameters; for example, 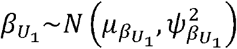 where 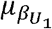 is the analyst’s best guess of the true value of 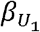 (e.g., a result from a published paper) and 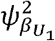 represents the analyst’s uncertainty about 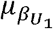. The bias model is estimated using Markov Chain Monte Carlo (MCMC) estimation, specifically Gibbs sampling implemented by JAGS (version 4.3.0 or later) [25]. The conditional posterior distribution of *β* _*X*_ is derived from the joint posterior distribution of all unknown quantities (i.e., all parameters of bias model eq:1 and the missing values of *U*).

Table 1 summarises the key features of *unmconf* as described in Hebdon et al [23]. However, R package *unmconf* can fit a greater range of bias models. In the simulation study and applied example below, we show that *unmconf* can fit our proposed bias model and is also applicable when exposure *X* is categorical.

### 2.3 Monte Carlo QBA qbaconfound

We propose a Monte Carlo QBA in which the bias model has the flexible features of *unmconf* (e.g., appliable to a range of substantive analyses and allows for correlations between *U* and *C*, and between *U*_1_ and *U*_2_) but with substantially fewer bias parameters. Our proposed Monte Carlo QBA will be implemented as Stata command and R package called *qbaconfound*. Without loss of generality, we describe the bias model of *qbaconfound* with respect to *k* continuous unmeasured confounders, *U*_1_, …, *U*_*k*_, and *p*-*k* binary unmeasured confounders, *U*_*k* +1_, ‥, *U*_*p*_, where (*U*_*j*_ = 1) = *π* _*j*_ for *j*= *k* + 1, …, *p*.

The *qbaconfound* bias model does not model *U* directly but instead models the part of *U* uncorrelated with *C* (i.e., the part of *U* not explained by *C*). This avoids having to specify the relationships between *U* and *C* whilst still allowing *U* and *C* to be correlated. Let *Ũ*_*j*_ denote the part of *U*_*j*_ that is uncorrelated with measured confounders *C* (*j* = 1, …, *p*) and *Ũ* = (*Ũ* _1_, ‥, *Ũ* _*p*_). We can think of each *Ũ* _*j*_ as the residuals from fitting regression *U*_*j*_ |c. We shall refer to *Ũ* as proxy variables for the unmeasured confounders *U*. Note that *Ũ* _1_, ‥, *Ũ* _*p*_ may still be correlated with each other; that is, if *U*_*j*_ is correlated with 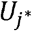 independently of *C* (i.e., 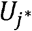 explains a part of *U*_*j*_ not explained by *C*, and vice versa) then *Ũ* _*j*_ and 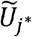 will also be correlated (*j, j*^*^ = 1, …, *p* and *j* ≠ *j*^*^). For *k* continuous unmeasured confounders and *p*−*k* binary unmeasured confounders, the bias model of *qbaconfound* is:

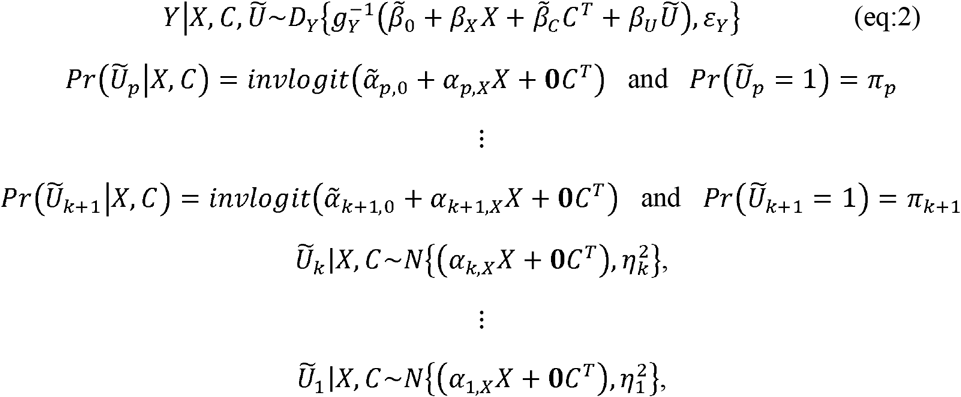

where **0** denotes a 1 × *q* vector of zeros, and the bias parameters are defined with respect to the unmeasured confounders *U* instead of their proxy variables *Ũ* ; that is, *β* _*U*_ denotes the relationship between *Y* and *U* (given *X* and *C*), 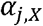 denotes the relationship between *U*_*j*_ and *X* (given *C*), (for continuous *U*_*j*_) 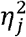 denotes the residual variance from linear regression *U*_*j*_|*X, C*, and (for binary *U*_*j*_) *π* _*j*_ is the marginal prevalence of binary *U*_*j*_. Note that for each binary *U*_*j*_, the analyst provides information about its marginal prevalence, *π* _*j*_, instead of the intercept term, 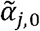,. The rationale is that a marginal prevalence is easier to comprehend and more readily reported in the published literature than an intercept of a logistic regression. A numerical approximation approach is used to derive the value of 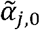, for given values of *π* _*j*_ and *α* _*j,X*_.

Given prespecified values of the bias parameters, the remaining parameters of the *qbaconfound* bias model (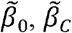 and ℰ_*Y*_), are estimated as follows: Step 1, for each *U*_*j*_ in turn, simulate the corresponding *Ũ* _*j*_ (e.g.,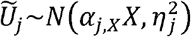) using information about the relationship between X and *U*_*j*_ given *C*. Step 2, fit regression *Y*| *X, C, Ũ* to the observed data (*Y, X*, and *C*) and the simulated *Ũ* = (*Ũ*_1_, …, *Ũ*_*p*_) using maximum likelihood estimation, with the coefficients of *Ũ* constrained to equal *β* _*U*_ (i.e., the coefficients of *U* from the substantive analysis).

Noteworthy points about the bias model are: (1) The coefficient of *X* from regression *Y*| *X, C, Ũ* is the exposure effect, *β* _*X*_. Supplementary section 2 gives a full explanation for this statement based on the principles of partitioning the variance of the dependent variable in a multiple regression [26]. Briefly, to estimate _X_ (from regression *Y*|*X, C, Ũ*), *Ũ* must be a good proxy for *U* with respect to its confounding effects on the *Y*-*X* association. First, by constraining the coefficient of *Ũ* to *β* _*U*_, the relationship between *Y* and *Ũ* (given *X* and *C*) is fixed to be the same as the relationship between *Y* and *U* (given *X* and *C*). Second, *Ũ* is simulated using information about the relationship between X and *U* given *C*. Therefore, the shared variance of *Y* between X and *Ũ* (i.e., the covariance) is fixed to be the same as the covariance between *X* and *U*. (2) Conversely, the coefficient of *C* (of regression *Y*| *X, C, Ũ*) differs from that of the substantive analysis because *Ũ* represents only the part of *U* uncorrelated with *C* and so *Ũ* does not account for the shared variance of *Y* between *U* and *C*.(3)Eq:2 does not model the relationships between *Ũ* _1_, ‥, *Ũ* _*p*_ and equally does not assume independence between *Ũ* _1_, …, *Ũ* _*p*_|*X, C* (and thus between *U*_1_, …, *U*_*p*_|*X, C*). Since the relationship between *Y* and *Ũ* (given *X* and *C*) is constrained to be *β*_*U*_ (i.e., not estimated), simulation of *Ũ* can ignore any shared variance of *Y* between the unmeasured confounders. (4) The number of bias parameters does not depend on the number of measured confounders. Therefore, for one binary and one continuous unmeasured confounder and 10 measured confounders, the *qbaconfound* bias model contains 6 bias parameters which is substantially lower than the 28 bias parameters of the *unmconf* bias model.

We propose conducting a Monte Carlo QBA using the *qbaconfound* bias model which requires specification of prior distributions for the bias parameters only. These are informative prior distributions, specifically a univariate normal distribution for each coefficient, and uniform distributions for the standard deviations and marginal prevalences. As described in section 2.2, external information is used to specify the values of the hyperparameters. The Monte Carlo procedure generates a frequency distribution of bias-adjusted estimates of *β*_*X*_ by repeatedly carrying out the following steps *L* (*L*> 1) times [11]: for *l*= 1, ‥, *L*

i. Randomly draw a value for each bias parameter from its prior distribution. Let 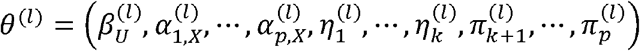 denote the drawn values for step *l*. For each binary *Ũ*_*j*_, derive intercept 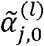 from drawn values 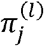 and 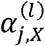.
ii. Fit the *qbaconfound* bias model to (*Y, X, C*), fixing the bias parameters to *θ*^(*l*)^. Step 1. Simulate 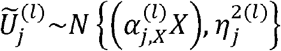 for *j* = 1, …, *k* and simulate

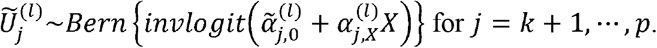

Step 2. For 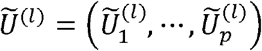, fit regression *Y*| *X, C, Ũ* ^(*l*)^ with the coefficients of Ũ ^(*l*)^ constrained to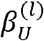. Let 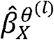 and 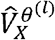 denote the point estimate and corresponding variance of *β*_*X*_, respectively.
iii. Incorporate random sampling error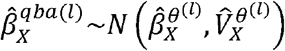.

After *L* replications, we compute the median, 2.5^th^ and 97.5^th^ percentiles of the frequency distribution 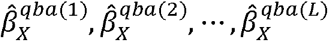 to obtain the Monte Carlo point and interval estimates of *β*_*X*_.

## 3. Simulation studies

We describe two simulation studies to evaluate the performance of our proposed Monte Carlo QBA, *qbaconfound*. Simulation study I compares Bayesian and Monte Carlo QBAs using the *qbaconfound* bias model (eq:2). Note that the Bayesian QBA was implemented using R package *unmconf*, and we did not compare the *qbaconfound* bias model to the bias model of *unmconf*. Simulation study II evaluates the *qbaconfound* bias model for differing levels of correlation between *C* and *U*, and, where relevant, between *U*_1_ and *U*_2_. In both simulation studies, we consider two study scenarios. In scenario A, we have binary *Y*, binary *X*, and a single continuous *U*= (*U*_1_), where the substantive analysis, *Y*|*X, C, U*, is a logistic regression. In scenario B, we have continuous *Y*, continuous *X*, and two continuous *U*= *U*_1_, *U*_2_, where *Y*|*X, C, U* is a linear regression. For both scenarios there are three measured confounders: *C* _*cat*_ = (*C* _*cat* 1_, *C* _*cat* 2_) is a 3-category variable with reference category 0 and dummy variables *C* _*cat* 1_ and *C* _*cat* 2_ (i.e., *C* _*cat j*_ =1 for category *j* and *C* _*cat*_ j =0 otherwise, for *j*= 1,2). *C*_*bin*_ is a binary variable, and *C*_*con*_ is a continuous variable. Therefore, *C*= (*C* _*cat* 1_, *C* _*cat* 2_, *C*_*bin*_, *C*_*con*_) consists of 4 variables (i.e., *q* = 4). For all combinations of the simulation settings, we generate 500 simulated datasets, each with 1,000 observations.

Below we first describe the data generation models and performance measures which are common to simulation studies I and II and then describe the specifics of each simulation study.

We used Stata version 18.0 [27] to generate the data and perform the Monte Carlo QBA and R 4.4.3 [28] to perform the Bayesian QBA, specifically using R package *unmconf* version 1.0.0 [17] implemented in JAGS 4.3.1 [25]. We modified the *unm_glm* function of package *unmconf* to correctly include the unmeasured confounding in the substantive analysis when only one unmeasured confounder is present and to enable our results to be reproducible.

### 3.1 Simulation study design

#### 3.1.1 Data generation

Data for simulation studies I and II were generated from the two data generating models described in supplementary section 3. Most of the parameters of the data generating models were based on an analysis of real data from the Barry Caerphilly Growth study [29, 30]. Briefly, both data generating models factorised the joint distribution of *X, Y, C*, and *U* as

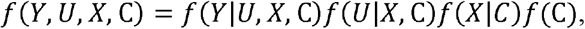

where *f* (·) denotes a probability or density function and *f* (*C*) = *f* (*C*_*con*_, *C*_*bin*_, *C*_*cat*_)= *f* (*C*_*con*_ |*C*_*bin*_, *C*_*cat*_)*f C*_*bin*_ |*C*_*cat*_)*f C*_*cat*_. Categorical *C*_*cat*_ was simulated from a (marginal) multinomial distribution, binary variables (e.g., *C*_*bin*_) from a logistic regression model, and continuous variables (e.g., *C*_*con*_) from a linear regression model.

Note that, in scenario A, the single continuous unmeasured confounder, was simulated from a linear regression model, and in scenario B the two continuous unmeasured confounders were simulated from a multivariate normal regression model. Note that the data were not generated such that the magnitude of the bias from omitting two unmeasured confounders (scenario B) was twice the magnitude of the bias from omitting a single unmeasured confounder (scenario A).

#### 3.1.2 Estimand and performance measures

The estimand of interest was exposure effect, *β*_*X*_. The true value of *β*_*X*_ was known because it was a parameter of the data generating models (null value for both scenarios). Performance measures of interest were bias and empirical standard error of estimates of *β*_*X*_, mean model-based standard errors of *β*_*X*_, and 95% coverage of interval estimates of *β* _*X*_. Performance measures were computed using Stata command *simsum* [31].

#### 3.1.3 Simulation study I

We compared our proposed Monte Carlo QBA to a Bayesian QBA using the same bias model; that is, the *qbaconfound* bias model of eq:2. The Monte Carlo QBA was applied with 100 Monte Carlo replications and the Bayesian QBA with 4,000 iterations of which 1,000 were burn-in iterations. (See supplementary sections 4 and 5 for further information on the choice of the number of Monte Carlo replications and iterations lengths, respectively).

We applied both QBAs with the same set of prior distributions for the bias parameters. We used normal distributions for coefficients 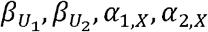 and uniform distributions for standard deviations 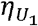 and 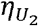. The mean hyperparameter of each normal prior was set to the true value of the corresponding bias parameter, and the variance hyperparameter and the upper and lower hyperparameters of each uniform distribution were set such that 95% of the values sampled directly from the prior were within ± *w* % of the true value. We applied both QBAs with two sets of priors that differed with respect to the level of certainty: very informative and informative priors where 95% of the directly sampled values were within ± 10% and ± 30% of the truth, respectively. Using the informative priors, we repeated these analyses with the uniform distributions replaced with Gamma distributions for precisions 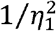 and 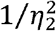. These conjugate priors are known to have good sampling efficiency [25]. For the Bayesian QBA, we specified vague priors for the non bias parameters, specifically 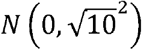 for coefficients and *Gamma* (0.001, 0.001) for the inverse of residual variances of linear regression models. All covariates were centred before applying the Bayesian QBA for sampling efficiency.

#### 3.1.4 Simulation study II

As discussed in section 2.3, the *qbaconfound* bias model does not explicitly model the associations between *U* and *C* nor between multiple unmeasured confounders (i.e., between *U*_1_ and *U*_2_). Neither does the model assume independence between *U* and *C* nor between *U*_1_ and *U*_2_. The aim of this simulation study was to evaluate the bias model when in truth (i) *U* and *C* were (marginally) independent (i.e., *f* (*U*|*X, C* = *f* (*U*|*X*) for all values of *C*) and *U*_1_ and *U*_2_ were (marginally) independent, and (ii) there were strong associations between *U* and *C* and between *U*_1_ and *U*_2_ (i.e., double the magnitudes of simulation study I).

We simulated data based on scenario A (logistic regression substantive analysis with binary *X, C*= *C*_*cat* 1_, *C*_*cat* 2_, *C*_*bin*_, *C*_*con*_, and continuous *U*= (*U*_1_)) and scenario B (linear regression substantive analysis with continuous *X, C*= (*C*_*cat* 1_, *C*_*cat* 2_, *C*_*bin*_, *C*_*con*_), and continuous *U*= (*U*_1_, *U*_2_). (See supplementary sections 3.1 to 3.3 for further details on the data generating models). For both scenarios, we analysed the data using the full substantive analysis model, *Y*|*X, C, U* (i.e., as if *U* were measured), the naïve analysis model, *Y*|*X, C*, and the Monte Carlo QBA, *qbaconfound*, with 100 replications and very informative priors for the bias parameters (as described in section 3.1).

### 3.5 Results

Figures 1 and 2 show the bias and coverage of estimating *β*_*X*_ using the full analysis model, naïve analysis model, and Monte Carlo or Bayesian QBA, from simulation studies I and II, respectively (detailed results reported in supplementary tables 4 and 5). In the absence of unmeasured confounding, all full analysis estimates of *β*_*X*_ were unbiased and confidence interval coverages were close to the nominal level. Note that the full analysis estimate for Scenario A was slightly biased due to our moderately small sample size [32]. As expected, given the design of the simulation studies, the naïve estimates were substantially biased with poor confidence interval coverage. We note that the exceptionally low coverage (3.4%) for scenario B was driven not just by the magnitude of bias but also by the distribution of deviations (i.e., differences in individual point estimates and true value). Note that for scenario A, the naïve analysis did not strictly estimate the intended estimand because *β*_*X*_ was a non-collapsible effect measure (i.e., log odds ratio). Below we describe the specific findings of simulations studies I and II.

**Figure 1.**
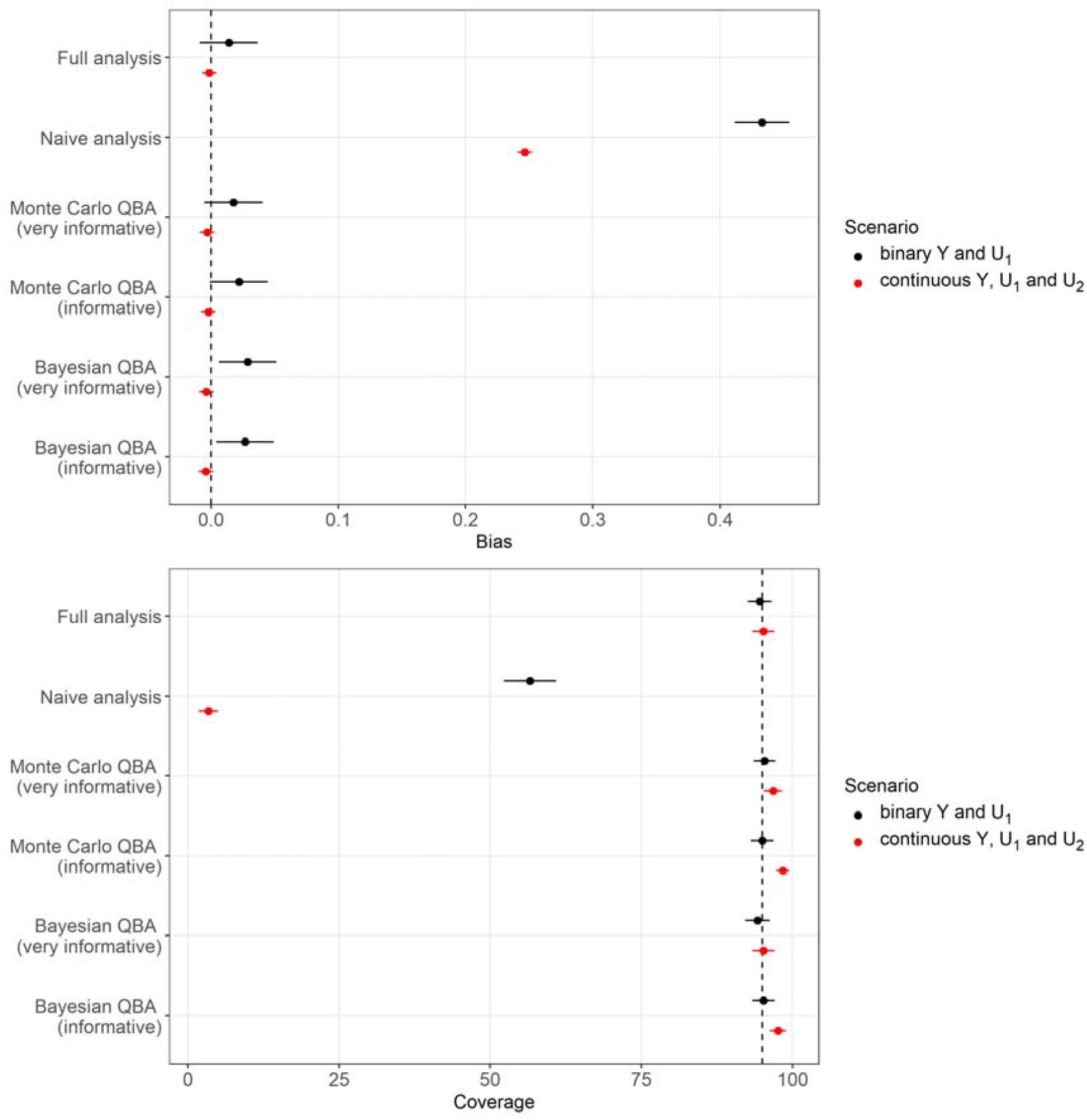
Bias and 95% coverage of point and interval estimates, respectively, of exposure effect *β*_*X*_ with a binary outcome and one unmeasured confounder, *U*_1_, (in black) and a continuous outcome with two unmeasured confounders, *U*_1_ and *U*_2_, (in red) where associations between *U* and *C* and between *U*_1_ and *U*_2_ were moderate. Error bars denote 95% Monte Carlo intervals. Vertical dashed line denotes zero bias (top) and nominal coverage (bottom).

**Figure 2.**
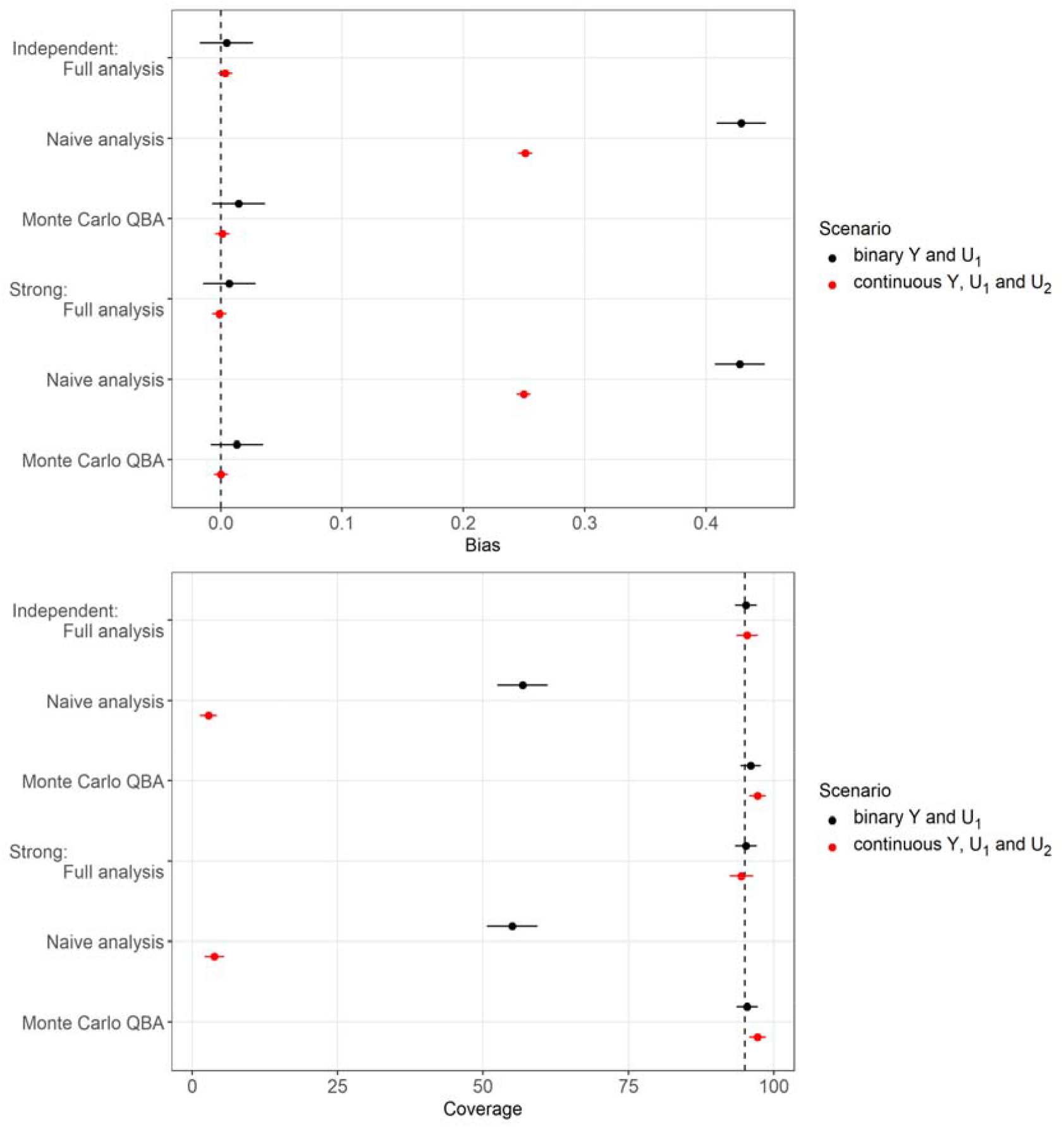
Bias and 95% coverage of point and interval estimates, respectively, of exposure effect *β*_*X*_ with a binary outcome and one unmeasured confounder, *U*_1_, (in black) and a continuous outcome with two unmeasured confounders, *U*_1_ and *U*_2_, (in red) where associations between *U* and *C* and between*U*_1_ and *U*_2_ were zero (independent) and strong. Error bars denote 95% Monte Carlo intervals. Vertical dashed line denotes zero bias (top) and nominal coverage (bottom).

#### 3.5.1 Simulation study I

For both scenarios, the Monte Carlo and Bayesian implementations performed equally well when using very informative and informative priors for the bias parameters. The point estimates were either unbiased (i.e., the 95% Monte Carlo interval for bias included 0) or had negligible levels of bias and interval coverages were at least nominal. We note that for scenario B (linear regression with two unmeasured confounders), there was evident overcoverage when using informative priors but only slight overcoverage when using very informative priors; clearly a reflection of the greater levels of uncertainty when using less informative priors. The fact that in scenario A (logistic regression with one unmeasured confounder) coverage levels were not as affected by the level of prior uncertainty was likely due to the fact that scenario B required double the number of bias parameters compared to scenario A.

The results were very similar when applying the Monte Carlo and Bayesian QBAs with a Gamma distribution for precisions 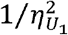 and 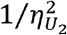 (results reported in supplementary tables 4 and 5). For both scenarios, the Bayesian QBA (with a total of 4,000 iterations) took slightly longer (up to 30 seconds per dataset) to run than the Monte Carlo QBA (approximately 1 second per dataset with 100 replications). All results except runtime were very similar when applying the Monte Carlo QBA with 400 and 10,000 replications (supplementary tables 2 and 3). As expected, the runtime increased with the number of replications.

#### 3.5.2 Simulation study II

Figure 2 shows that the performance of *qbaconfound* (Monte Carlo implementation of our proposed bias model) is unaffected by the level of the true associations between *U* and *C*, and between *U*_1_ and *U*_2_ in scenario B. The results were very similar to those of figure 1 where the point estimates were either unbiased or had negligible levels of bias, and interval coverages were nominal (detailed results in supplementary tables 6 and 7).

#### 3.5.3 Other results

Applying the Monte Carlo QBA in other settings resulted in unbiased (or negligible bias) of the point estimate of *β*_*X*_ and nominal interval coverages: logistic regression with a binary *X* and binary *U*= (*U*_1_) (supplementary table 8), linear regression with a continuous *X*, and binary *U*= (*U*_1_, *U*_2_) (supplementary table 9), linear regression with a categorical *X*, and a continuous and binary *U*= (*U*_1_, *U*_2_) (supplementary table 10), multinomial regression with a continuous *X* and continuous *U*= (*U*_1_) (supplementary table 11), and Cox proportional hazards regression with a continuous *X* and continuous *U*= (*U*_1_) (supplementary table 12).

## 4. Applied example

We illustrate the implementation of our Monte Carlo QBA method, *qbaconfound*, when the analysis is a linear regression with a categorical exposure *X* and two unmeasured confounders *U*= (*U*_1_, *U*_2_)where *U*_1_ is continuous and *U*_2_ is binary. As with our simulation study, we repeated the analysis as a Bayesian QBA implementing the same *qbaconfound* bias model using R package *unmconf*. We used the publicly accessible data from the 2015-2012 (2003-2004, 2005-2006, 2007-2008, 2009-2010, and 2011-2012) National Health and Nutrition Examination Survey (NHANES) study [33, 34, 35, 36, 37]. □ The NHANES study consists of a series of health and nutrition surveys conducted by the National Center for Health Statistics. Based on an analysis reported in [38], we examined the association of alcohol drinking quantity with waist circumference (cm) among 7477 men between 20 and 79 years old who are currently not on a medical or weight-gain/loss diet. Note that [38] used sampling weights to reflect the complex sampling design of the NHANES survey data. However, we did not conduct a weighted analysis because we have not evaluated *qbaconfound* in this setting and R package *unmconf* does not support the use of weights. Alcohol drinking quantity was first categorised into three levels: “never”, “former” and “current” drinkers. “Current” drinkers were further categorised into three levels based on the average number of drinks on a drinking day: those who drank 1 to 2 drinks (baseline), 3 to 4 drinks and at least 5 drinks. Measured covariates *C* were age, race/ethnicity, education, income, survey year of data collection, smoking status, whether day of dietary recall fell on a weekday or weekend on two recall days, physical activity level and dietary misreporting status (as defined in [38]). □ Continuous *U*_1_ was daily non-alcoholic energy intake (kcal) and *U*_2_ was marital status (0 currently in a relationship, 1 currently single). Note that confounders *U*_1_ and *U*_2_ were measured by the NHANES study and were included in the analyses of [38]. For the purposes of illustrating a QBA, we treat *U*_1_ and *U*_2_ as unmeasured. Also, as this is an illustrative example, we have ignored other potential sources of bias (such as missing data), and we restricted our analyses to participants with complete data on *Y, X, C* and *U*= (*U*_1_, *U*_2_).

The *qbaconfound* bias model is given by eq:2 with a normal distribution for *D*_*Y*_{·}, an identity function for 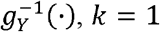 and *p* = 2. The priors for the bias parameters were normal distributions for regression coefficients 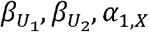, and α _2,*X*_ and uniform distributions for residual standard deviation 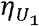 and marginal prevalence *π* _2_ (see supplementary table 13 for more details). Note that for the Bayesian QBA, we had to specify a prior (a normal distribution) for the intercept term 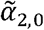, instead of *π*_2_. This was due to the restrictions of the *unmconf* R package. We repeated the Monte Carlo QBA using the same prior for 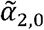, (which replaced the prior for *π*_2_). The hyperparameters of the bias parameters’ priors were set to values from analysing the observed data (since in reality *U*_1_ and *U*_2_ were measured; see supplementary table 13 for further details). For example, for prior 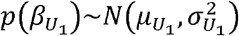 we set 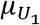 and 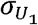 to equal the point estimate and standard error of the coefficient of *U*_1_ from linear regression *Y*|*X, C, U*_1_, *U*_2_. For the Bayesian QBA, we specified vague priors for the non bias parameters, specifically *N* (0, 10^2^) for the coefficients (e.g., *β*_*X*_) and *Gamma* (0.001, 0.001) for the inverse of residual variance (i.e., precision) 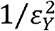. The Bayesian QBA was applied with a chain of 22,000 iterations of which 2,000 were burn-in iterations, and the Monte Carlo QBA was applied with 10,000 replications.

We used Stata version 18.0 [27] to perform the Monte Carlo QBA and R 4.4.3 [28] to generate the NHANES analysis sample and perform the Bayesian QBA using R package *unmconf* version 1.0.0 [17] implemented in JAGS 4.3.1 [25].

Figure 3 shows the results for the *β*_*X*_ estimates, using the full analysis model, naïve analysis model, and Monte Carlo and Bayesian QBAs (full details in supplementary table 14). All analyses suggest that former drinkers and those who drank at least 5 drinks per drinking day have a significantly wider waist circumference on average compared to those who only drank 1 to 2 drinks. The full and naïve analyses produced different point estimates but there was substantial overlap between the confidence intervals due to large standard errors. The Monte Carlo and Bayesian QBAs gave similar results that are close to the full analysis.

**Figure 3.**
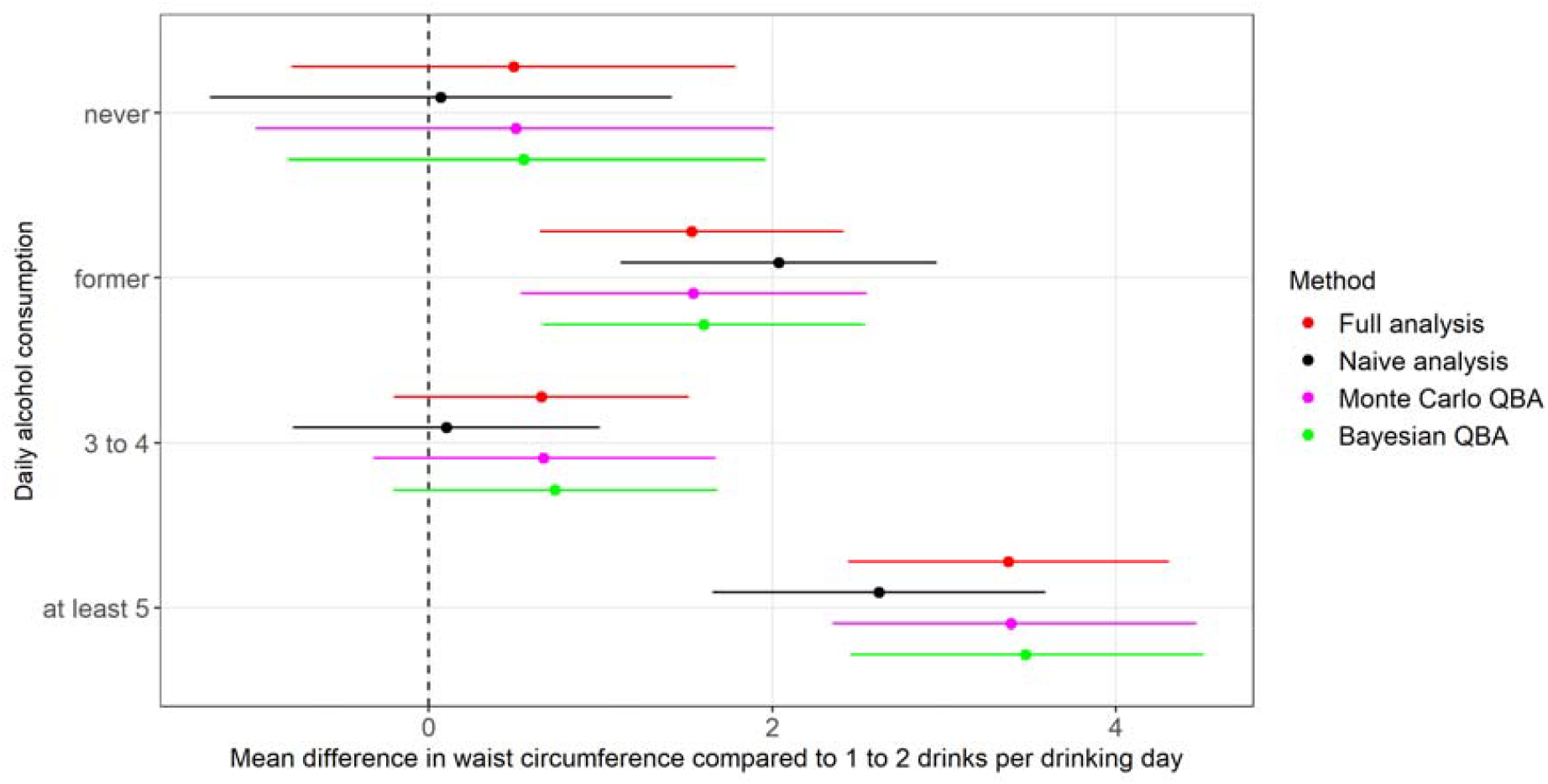
Forest plot of the results for exposure effects, *β*_*X*_, for the four non-baseline levels of alcohol drinking quantity estimated by the full analysis, naïve analysis, Monte Carlo^$$^, and Bayesian quantitative bias analyses (QBAs). Dashed line denotes no difference to those who drank 1 to 2 drinks per drinking day (baseline). $$ Mote Carlo QBA using prior *p* (π_2_)

## 5. Discussion

We have proposed a flexible Monte Carlo QBA approach to unmeasured confounding, *qbaconfound*, that minimises the burden placed on the user by keeping the number of bias parameters to a minimum and avoiding the need for specialist knowledge about Bayesian inference or Bayesian software. The *qbaconfound* bias model is applicable for a wide range of regression analyses (GLMs and Cox PH regression with binary, continuous, or categorical exposures) with one or multiple (two or more) unmeasured confounders, and allows for correlation between all confounders (i.e., between measured and unmeasured confounders, and between multiple unmeasured confounders). Our extensive simulation study demonstrated that *qbaconfound*, with informative priors for the bias parameters, gives unbiased (or minimally biased) estimates of the exposure effect, and with nominal interval coverage, in a wide range of scenarios. Also, we have shown that the Monte Carlo QBA (using our proposed bias model) performed as well as the Bayesian QBA, which is in keeping with a previous study comparing these two approaches in the context of nonignorably missing data [6].

There are two key differences between *qbaconfound* and *unmconf*. First, unlike with *unmconf*, the number of bias parameters of *qbaconfound* does not depend on the number of measured confounders. This is key since analyses of observational studies tend to involve many measured confounders. Second, *qbaconfound* implements a Monte Carlo QBA and *unmconf* implements a Bayesian QBA. Clearly, *qbaconfound* is advantageous over *unmconf* for studies with no access to suitable validation data or for users who lack the specialist knowledge to carry out (or are daunted by) Bayesian inference. However, for those who prefer the more principled Bayesian approach, then the *qbaconfound* bias model can be implemented as a Bayesian QBA using the *unmconf* R package (as illustrated in our simulation study and applied example). Lastly, the *unmconf* bias model is preferable to the *qbaconfound* bias model when the user is interested in all parameters of the substantive analysis (i.e., not simply the exposure effect). Unlike *qbaconfound, uncomf* provides estimates of all parameters of the user’s substantive analysis.

A key strength of our simulation study was our evaluation of *qbaconfound* QBA in a wide range of scenarios (with the majority of the results reported in the supplementary materials). However, inevitably we were not able to explore all possible analysis scenarios (such as Cox PH regression with multiple unmeasured confounders) in a single paper. We welcome future studies to investigate other scenarios. Another limitation is that our simulation study did not compare *qbaconfound* QBA to the *unmconf* QBA. Our rationale was that our primary objective was to conduct a thorough evaluation of our proposed *qbaconfound* QBA. Also, *qbaconfound* QBA is not intended as a replacement for *unmconf* QBA but instead as an alternative for the situation where a study which does not have access to validation data. Our third limitation is that we did not compare the Monte Carlo approach to the Bayesian approach when using vague priors for the bias parameters. Previous studies have shown that in this setting the results of the Monte Carlo approach were comparable to those of the naïve analysis whilst the results of the Bayesian approach were biased but to a lesser extent because some information about the bias parameters was gained from combining the prior with the likelihood of the observed data [6, 11]. Our rationale for not comparing the two approaches with vague priors is that it is not practicable to conduct a probabilistic bias analysis (either Bayesian or Monte Carlo) without some information about the bias parameters (i.e., unable to make conclusions due to excessively wide interval estimates). In the absence of any information about the bias parameters, a deterministic QBA could be conducted to at least determine if the study conclusions are sensitive or robust to unmeasured confounding. However, if there is evidence of sensitivity then a deterministic QBA is still unable to quantify the likely magnitude of the potential bias due to the lack of information about plausible values for the bias parameters.

We have proposed a simple to implement Monte Carlo QBA approach that is applicable in a wide range of substantive analyses, with one or multiple unmeasured confounders and requires only essential information about the unmeasured confounders. Given that validation data can be difficult to source, we consider *qbaconfound* an essential tool for performing a probabilistic QBA for unmeasured confounding.

## Data Availability

Barry Caerphilly Growth study: Ethical approval for the study was given by the Bro Taf Health Authority Local Research Ethics Committee. Data from the NHANES study is publicly available https://wwwn.cdc.gov/nchs/nhanes/default.aspx.

