## Supplementary materials for "*qbaconfound*: A flexible Monte Carlo probabilistic bias analysis for unmeasured confounding"

**bias analysis for unmeasured confounding**

### **1. Notation**

Supplementary table 1: Description of the notation used in the main paper

| **Symbol** | **Description** |
| --- | --- |
| $Y$ | Outcome of interest: continuous, binary, nominal, ordinal, or $Y=\left( S,D \right)$ for survival time $S$ and event indicator $D$. |
| $X$ | Exposure of interest. |
| $q^{*}$ | Number of measured confounders. |
| $C_{j}$ | Variable corresponding to a measured confounder such as a linear, or polynomial term of a continuous measured confounder, a binary measured confounder, a dummy variable of a categorical measured confounder, or an interaction term between two measured confounders, for $j=1,\cdots,q$ and $q\geq q^{*}$. |
| $C$ | q-vector of variables corresponding to the measured confounders; $C=\left( C_{1},\cdots,C_{q} \right)$ for $q\geq q^{*}$. |
| $p$ | Number of unmeasured confounders. |
| $U_{j}$ | Binary or continuous unmeasured confounder for $j=1,\cdots,p$. |
| $U$ | $p$-vector of unmeasured confounders; $U=\left( U_{1},\cdots,U_{p} \right)$. |
| $Y\left\vert X,C,U \right.$ | Substantive analysis of interest. |
| $\beta_{X}$ | Exposure effect of interest. |
| $\beta_{C}$ | q-vector of coefficients corresponding to the measured confounders; $\beta_{C}=\left( \beta_{C_{1}},\cdots,\beta_{C_{q}} \right)$ where $\beta_{C_{j}}$ is the coefficient of $C_{j}$ in the substantive analysis (for $j=1,\cdots,q$). |
| $\beta_{U}$ | $p$-vector of coefficients of corresponding to the unmeasured confounders; $\beta_{U}=\left( \beta_{U_{1}},\cdots,\beta_{U_{p}} \right)$ where $\beta_{U_{j}}$ is the coefficient of $U_{j}$ in the substantive analysis ($j=1,\cdots,p$). Also, bias parameter of the *umnconf* and *qbaconfound* bias models. |
| $p\left( \cdot\right)$ | Prior probability distribution. |
| $D_{Y}\left\{ \cdot\right\}$ | Response distribution of generalised linear model for $Y$. |
| $g_{Y}^{-1}\left( \cdot\right)$ | Link function of generalised linear model for $Y$. |
| $D_{U_{j}}\left\{ \cdot\right\}$ | Response distribution of generalised linear model for $U_{j}$ $\left( j=1,\cdots,p \right)$. |
| $g_{U_{j}}^{-1}\left( \cdot\right)$ | Link function of generalised linear model for $U_{j}$ $\left( j=1,\cdots,p \right)$. |
| $\tilde{U}_{j}$ | Part of $U_{j}$ uncorrelated with $C$ for $j=1,\cdots,p$. |
| $\tilde{U}$ | $p$-vector $\tilde{U}=\left( \tilde{U}_{1},\cdots,\tilde{U}_{p} \right)$. |
| $\boldsymbol{0}$ | $1\times q$ vector of zeros. |
| $\alpha_{j,X}$ | Coefficient of $X$ in regression $U_{j}\left\vert X,C \right.$ and bias parameter of *qbaconfound* for $j=1,\ldots,p$. |
| $\eta_{j}^{2}$ | Residual variance of linear regression $U_{j}\left\vert X,C \right.$ and linear regression $\tilde{U}_{j}\left\vert X,C \right.$; bias parameter of *qbaconfound* for continuous $U_{j}$ ($j=1,\ldots,p$). |
| $\pi_{j}$ | Marginal prevalence of binary $U_{j} (j=1,\ldots,p).$ |
| $m$ | Number of replications used to perform the Monte Carlo probabilistic bias analysis |

### **2. Derivation of the bias model of *qbaconfound***

In this section, we explain how we derive our bias model (eq:S2) from Hebdon et al’s specification in eq:S1 [1]. Without any loss of generality, we consider a scenario in which there is a single measured and a single unmeasured confounder, $C=\left( C_{1} \right)$ and $U=\left( U_{1} \right)$, and the bias model consists of linear regression models. Note that the arguments below are based on variance partitioning of the linear predictor of a GLM which apply to all generalised linear models and an unspecified number of confounding variables (i.e., covariates of a regression).

Hebdon et al model the conditional joint distribution of $Y$ and $U$ given $X$ and $C$ using linear regressions:

$Y\left| X,C,U\sim N\left\{ \left( \beta_{0}+\beta_{X}X+\beta_{C}C+\beta_{U}U \right),\sigma^{2} \right\} \right.$, (eq:S1)

$U\left| X,C\sim\right.N\left\{ \left( \alpha_{0}+\alpha_{X}X+\alpha_{C}C \right),\eta^{2} \right\},$

where $Y\left| X,C,U \right.$ is the substantive analysis, $\beta_{X}$ is the exposure effect of interest, and $\beta_{U},\alpha_{0},\alpha_{X},\alpha_{C}$ and $\eta^{2}$ are the bias parameters.

In multiple regression, each covariate coefficient is interpreted as the unique effect of that covariate on the dependent variable (i.e., expected change in the dependent variable per one-unit change of the covariate whilst keeping the other covariates constant). We can think of the unique effect of, say, covariate $X$ in terms of the proportion of the variance of the dependent variable that is uniquely explained by $X$. Supplementary figure S1 illustrates the partitioning of the variance in multiple regressions $Y\left| X,C,U \right.$ and $U\left| X,C \right.$, where the variance of each variable is represented by its own ellipse and overlap between the ellipses represents shared variance (i.e., covariance) between the variables. For example, the proportion of the variance of $Y$ not explained by $X,C$, or $U$ is labelled as region $A_{Y}$ and the covariance between $Y$ and $U$ is labelled as region $A_{yu}$. In principle, $\beta_{X}$ corresponds to the unique proportion of the variance of $Y$ solely explained by $X$, region $A_{yx}$. Similarly, $\beta_{C}$ and $\beta_{U}$ correspond to the unique proportions labelled as regions $A_{yc}$ and $A_{yu}$, respectively. However, regions $A_{yxu},A_{yxc},A_{ycu}, A_{yxc},A_{ycu},$ and $A_{yxuc}$ cannot be attributed to a single covariate and indicate the redundancy of the model due to the shared variance of $Y$ among the covariates (i.e., known as collinearity). When covariates are highly collinear, the standard errors of the coefficients are often large and estimates of the coefficients can be highly variable between different studies [2]. Therefore, alternative methods to multiple regression are recommended when there is high collinearity. We assume that regressions $Y\left| X,C,U \right.$ and $U\left| X,C \right.$ are not prone to high levels of collinearity and if $U$ had been measured then the parameters of these regressions could be reliably estimated [2].

For regression $U\left| X,C \right.$ the variance of $U$ is partitioned into regions $A_{u}+A_{yu}$, $A_{xu}+A_{yxu}$, $A_{uc}+A_{yuc}$, and $A_{xuc}+A_{yxuc}$. Note that these regions are expressed in terms of the partitioning regions of regression $Y\left| X,C,U \right.$. For example, since $Y$ is not a covariate of regression $U\left| X,C \right.$ then the unique effect of $X$ (i.e., coefficient $\alpha_{X}$) is the proportion of the variance of $U$ that is explained by $X$ only and by $X$ and $Y$ which is represented by the sum of regions $A_{xu}+A_{yxu}$. The residual variance of $U$ (i.e., $\eta^{2}$) is represented by the sum of regions $A_{u}+A_{yu}$.

Let $\tilde{U}$ denote the part of $U$ uncorrelated with $C$, where $\tilde{U}$ is centred such that the intercept of $\tilde{U}\left| X,C \right.$ is zero. Replacing $U$ with $\tilde{U}$, the bias model becomes

$Y\left| X,C,\tilde{U}\sim N\left\{ \left( \tilde{\beta}_{0}+\beta_{X}X+\tilde{\beta}_{C}C+\beta_{U}\tilde{U} \right),\sigma^{2} \right\} \right.$,	(eq:S2)

$\tilde{U}\left| X,C\sim\right.N\left\{ \left( \alpha_{X}X \right),\eta^{2} \right\}$.

Note that for regression $Y\left| X,C,\tilde{U} \right.$ the coefficient of $X$ is the exposure effect of interest, $\beta_{X}$, and the coefficient of $\tilde{U}$ is bias parameter $\beta_{U}$. To understand why, consider supplementary figure S2 which shows the partitioning of the variances in regressions $Y\left| X,C,\tilde{U} \right.$ (left-hand subfigure) and $\tilde{U}\left| X,C \right.$ (right-hand subfigure) expressed in terms of the partitioning regions of the substantive analysis, $Y\left| X,C,U \right.$. We can see that the proportion of the variance of $Y$ that is uniquely explained by $X$ is the same for regressions $Y\left| X,C,U \right.$ and regression $Y\left| X,C,\tilde{U} \right.$ (i.e., region $A_{yx}$); hence, the coefficient of $X$ is the same in both regressions.

Similarly, the coefficients of $U$ and $\tilde{U}$ in regressions $Y\left| X,C,U \right.$ and $Y\left| X,C,\tilde{U} \right.$, respectively, are the same (i.e., $\beta_{U}$) because $U$ and $\tilde{U}$ uniquely explain the same proportion of the variance of $Y$ (i.e., region $A_{yu}$). Conversely, the coefficient of $C$ differs between the two regressions because the proportion of the variance of $Y$ that is uniquely explained by $C$ is region $A_{yc}$ in regression $Y\left| X,C,U \right.$ but it is the sum of regions $A_{yc}+A_{ycu}$ for regression $Y\left| X,C,\tilde{U} \right..$

We can follow a similar argument for multiple regression $\tilde{U}\left| X,C \right.$ to show that the coefficient of $X$ is $\alpha_{X}$ and the residual variance is $\eta^{2}$ (i.e., by comparing the partitioning of the variances of $U$ and $\tilde{U}$ according to regressions $U\left| X,C \right.$ and $\tilde{U}\left| X,C \right.$, respectively). Here, the coefficient of $C$ is 0 because $C$ does not explain any of the variance of $\tilde{U}$.

| 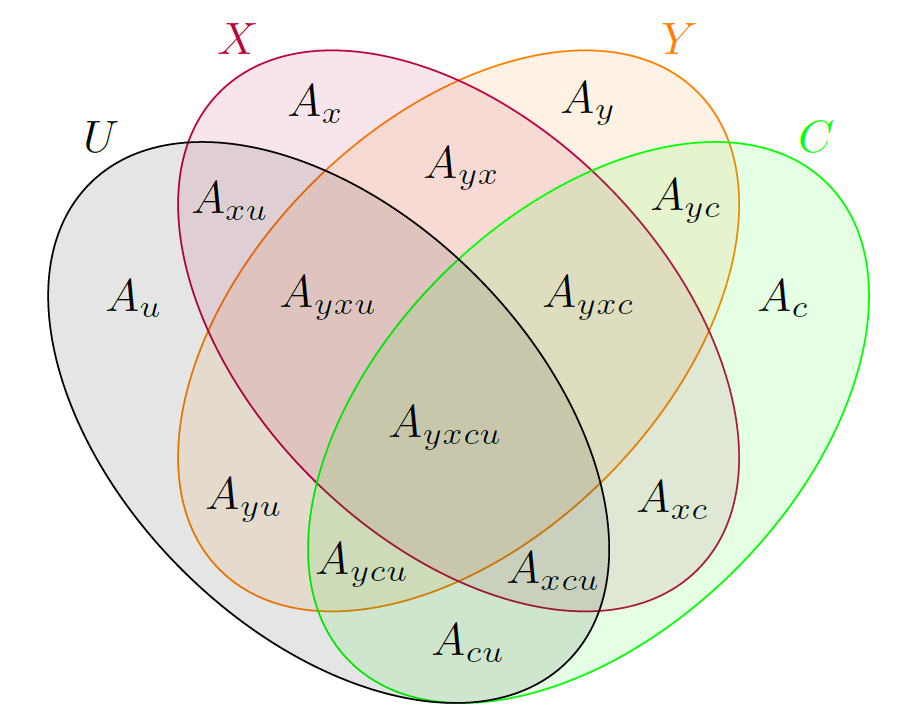 | 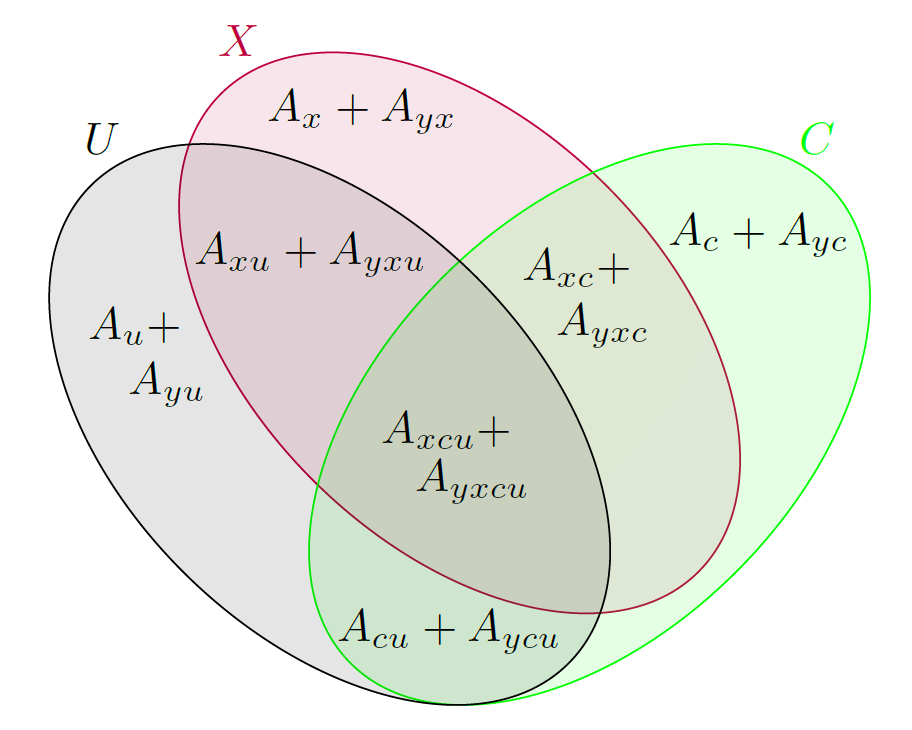 |
| --- | --- |
| Figure S1: Partitioning the variance in multiple regression models $Y\left\vert X,C,U \right.$ (left) and $U\left\vert X,C \right.$ (right). Each ellipse represents the variance of a variable ($Y,X,C,$ or $U$) and overlapping regions represent covariance between variables. | |

| 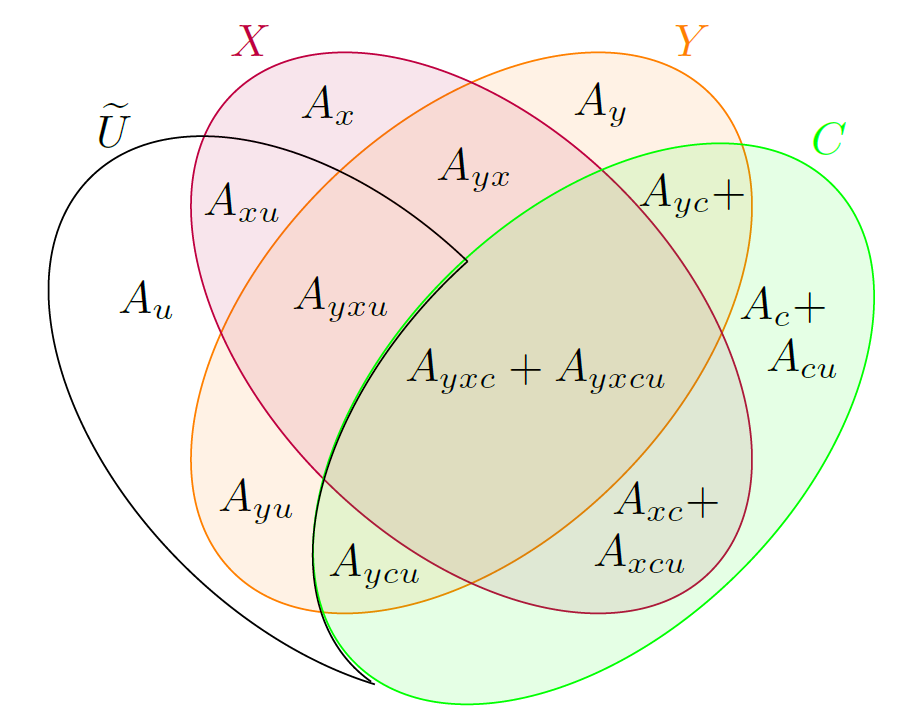 | 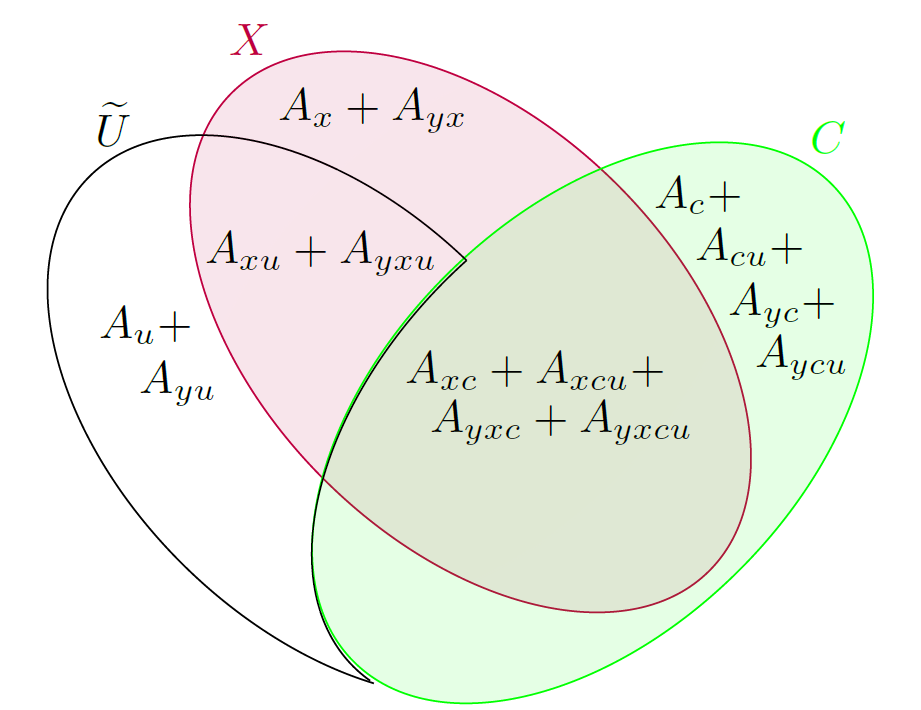 |
| --- | --- |
| Figure S2: Partitioning the variance in multiple regression $Y\left\vert X,C,\tilde{U} \right.$ (left) and $\tilde{U}\left\vert X,C \right.$ (right) Each ellipse (or portion of an ellipse for $\tilde{U}$) represents the variance of a variable and overlapping regions represent covariance between variables. | |

### **3. Simulation study: data generating models of scenarios A and B**

#### 3.1 Simulating measured confounders $C$

For both scenarios there are three measured confounders: $C_{cat}=\left( C_{cat1},C_{cat2} \right)$ is a 3-category variable with reference category 0 and dummy variables $C_{cat1}$ and $C_{cat2}$ (i.e., $C_{catj}=1$ for category $j$ and $C_{catj}=0$ otherwise, for $j=1,2$). $C_{bin}$ is a binary variable, and $C_{con}$ is a continuous variable. Data on $C_{cat}, C_{bin},$ and $C_{con}$ were simulated according to factorisation:

$f\left( C_{con},C_{bin},C_{cat} \right)=f\left( C_{con} | C_{bin},C_{cat} \right)f\left( C_{bin} | C_{cat} \right)f\left( C_{cat} \right)$,

where $f\left( \cdot\right)$ denotes a probability or density function. The specific details are:

$C_{cat} \sim multinomial\left( p_{0}=0.1261, p_{1}=0.7477, p_{2}=0.1261 \right)$,

$Pr\left( C_{bin}=1 \right)=expit\left\{ 0.2624-0.3651 C_{cat1}-0.4075 C_{cat2} \right\}$,

$C_{con}\sim N\left( 2.916+0.5791 C_{cat1}+0.6344C_{cat2}-0.1057 C_{bin},{0.4495}^{2} \right)$,

where $p_{j}=Pr\left( C_{cat}=j \right)$ for $j=0,1,2$.

#### 3.2 Scenario A: data generating model for $X$, $Y$, and $U=\left( U_{1} \right)$

Given data on measured confounders $C=\left( C_{con},C_{bin},C_{cat} \right)$, binary $Y$, binary $X$, and a single continuous $U=\left( U_{1} \right)$ were simulated as follows:

$Pr\left( X=1 \right)=expit\left\{ -4.819-0.3487C_{cat1}-0.7207 C_{cat2}+0.4609 C_{bin}+1.024 C_{con} \right\}$,

$U\sim N\left( 49.86-\left( \zeta\times0.9003 \right)C_{cat1}-\left( \zeta\times0.01227 \right)C_{cat2}+\left( \zeta\times0.2349 \right)C_{bin}+\left( \zeta\times3.296 \right)C_{con}+\left( \zeta\times6.636 \right)X, {9.523}^{2} \right)$,

$Pr\left( Y=1 \right)=expit\left\{ -2.370+0.1753 C_{cat1}+0.04492C_{cat2}-0.3559C_{bin}-0.009095C_{con}+0 X+0.06588 U \right\}$,

where $\zeta$ influences the magnitude of the associations between $U$ and $C$. We considered three levels of $\zeta$: $\zeta=0$ corresponds to independence between $U$ and $C$ given $X$, $\zeta=1$ corresponds to setting the associations between $U$ and $C$ to the observed associations from a real study, and $\zeta=2$ corresponds to strong (i.e., double the observed) associations between $U$ and $C$. In simulation study I we set $\zeta=1$ and in simulation study II we repeated the study for $\zeta=0$ and $\zeta=2$.

#### 3.3 Scenario B: data generating model for $X$, $Y$, and $U=\left( U_{1},U_{2} \right)$

Given data on measured confounders $C=\left( {C_{con},C}_{bin},C_{cat} \right)$, continuous $Y$, continuous $X$, and two continuous $U=\left( U_{1},U_{2} \right)$ were simulated as follows:

$X\left| C \right.\sim N\left( 13.41-0.1988 C_{cat1}-0.3895 C_{cat2}+0.1442 C_{bin}+1.719 C_{con}, {2.176}^{2} \right)$,

$\left. \begin{matrix} U_{1} \\ U_{2} \end{matrix} \right|X,C\sim N\left( \left[ \begin{matrix} \mu_{U_{1}} \\ \mu_{U_{2}} \end{matrix} \right],\left[ \begin{matrix} {9.394}^{2} & \zeta\times4.502 \\ \zeta\times4.502 & {10.30}^{2} \end{matrix} \right] \right)$,

$Y\left| X,C,U \right.\sim N\left( 8.534+0.5421C_{cat1}+0.5630C_{cat2}-0.2531C_{bin}-0.1783 C_{con}+0\times X+0.1299\times U_{1}+0.06747 U_{2}, {4.322}^{2} \right)$,

where

$\mu_{U_{1}}=37.91-\left( \zeta\times0.8979 \right) C_{cat1}-\left( \zeta\times0.1473 \right) C_{cat2}+\left( \zeta\times0.5159 \right)C_{bin}+\left( \zeta\times2.209 \right) C_{con}+1.260 X$,

$\mu_{U_{2}}=58.11-\left( \zeta\times0.4942 \right) C_{cat1}-\left( \zeta\times0.8886 \right) C_{cat2}+\left( \zeta\times0.05031 \right)C_{bin}+\left( \zeta\times0.1617 \right)C_{con}+1.281 X$,

and $\zeta$ influences the magnitude of the associations between $U$ and $C$, and between $U_{1}$ and $U_{2}$. In simulation study I, we set $\zeta=1$ (i.e., corresponds to setting the associations between $U$ and $C$, and between $U_{1}$ and $U_{2}$ to the observed associations from a real study), and in simulation study II we repeated the study for $\zeta=0$ (i.e., corresponds to independence between $U$ and $C$ given $X$, and between $U_{1}$ and $U_{2}$ given $X$ and $C$) and $\zeta=2$ (i.e., corresponds to strong (i.e., double the observed) associations between $U$ and $C$ and between $U_{1}$ and $U_{2})$.

### **4. Choosing the number of replications for the Monte Carlo QBA**

We chose a value for the number of replications, $m$, such that a repeat analysis of the same data would produce very similar results. Following the advice on choosing the number of imputations for multiple imputation [3] and the number of repetitions in a simulation study [4, 5], we selected a value for $m$ such that the Monte Carlo error of the bias-adjusted exposure effect point estimate, $MCE\left\{ \hat{\beta}_{X}^{MC} \right\}=\frac{SD\left\{ \hat{\beta}_{X}^{MC} \right\}}{\sqrt{m}}$, was approximately $r\%$ of the model standard error, $SE\left\{ \hat{\beta}_{X}^{MC} \right\}$. When $\hat{\beta}_{X}^{MC}$ is unbiased and its interval estimate is $\hat{\beta}_{X}^{MC}\pm1.96\times SE\left\{ \hat{\beta}_{X}^{MC} \right\}$, nominal coverage requires $SE\left\{ \hat{\beta}_{X}^{MC} \right\}\cong SD\left\{ \hat{\beta}_{X}^{MC} \right\}$. Setting $SE\left\{ \hat{\beta}_{X}^{MC} \right\}=SD\left\{ \hat{\beta}_{X}^{MC} \right\}$, $m$ is then simply:

$m=\left( \frac{SE\left\{ \hat{\beta}_{X}^{MC} \right\}}{\frac{r}{100}\times SE\left\{ \hat{\beta}_{X}^{MC} \right\}} \right)^{2}=\left( \frac{100}{r} \right)^{2}$.

Therefore, when $r=10\%, 5\%,$ and $1\%$ we require $m=100,$ $400$, and $10,00$0 replications, respectively.

Supplementary tables 2 and 3 show the results of applying the Monte Carlo approach with $m=100,$ $400$, and $10,000$ replications to simulated data of scenario A (binary $Y$, binary $X$, and continuous $U=\left( U_{1} \right))$ and scenario B (continuous $Y$, continuous $X$, and continuous $U=\left( U_{1},U_{2} \right)$), respectively. Except for mean runtime, the results for $m=100$ were very similar to those of $m=400$ and $m=10,000$. As expected, the runtime increases with the number of replications.

**Supplementary table 2:** Scenario A: Binary $Y$, binary $X$, and continuous $U=\left( U_{1} \right)$, summary of results for $\beta_{X}$ from the Monte Carlo quantitative bias analysis with differing number of replications, $m$: bias and empirical standard error of the point estimate, mean of model standard error, coverage of 95% interval estimate, and mean runtime. [95% Monte Carlo interval]

| **Prior** | $\boldsymbol{m}$ | **Bias** | **Empirical standard error** | **Mean**  **standard error** | **Coverage %** | **Mean runtime**  **in seconds** |
| --- | --- | --- | --- | --- | --- | --- |
| Very informative; Uniform prior for $\eta_{U}$ | 100 | 0.0175  [-0.00528, 0.0403] | 0.260  [0.244, 0.276] | 0.248  [0.246, 0.251] | 95.4  [93.6, 97.2] | 1.30  [1.29, 1.32] |
|  | 400 | 0.0197  [-0.00300, 0.0424] | 0.259  [0.243, 0.275] | 0.248  [0.246, 0.250] | 94.8  [92.9, 96.7] | 5.10  [5.07, 5.13] |
|  | 10,000 | 0.0188  [-.00375, 0.0414] | 0.258  [0.242, 0.274] | 0.249  [0.247, 0.250] | 94.4  [92.4, 96.4] | 121  [120, 122] |
| Informative; Uniform prior for $\eta_{U}$ | 100 | 0.0218  [-0.000839, 0.0445] | 0.259  [0.243, 0.275] | 0.263  [0.260, 0.265] | 95.0  [93.1, 96.9] | 1.32  [1.31, 1.33] |
|  | 400 | 0.0211  [-0.00145, 0.0437] | 0.257  [0.241, 0.273] | 0.263  [0.262, 0.265] | 95.4  [93.6, 97.2] | 5.18  [5.13, 5.22] |
|  | 10,000 | 0.0213  [-0.00131, 0.0439] | 0.258  [0.242, 0.274] | 0.264  [0.262, 0.265] | 95.4  [93.6, 97.2] | 118  [117, 118] |
| Informative; Gamma prior for $1/{\eta_{U}^{2}}$ | 100 | 0.0237  [0.000761, 0.0466] | 0.262  [0.245, 0.278] | 0.264  [0.262, 0.267] | 95.6  [93.8, 97.4] | 1.25  [1.24, 1.25] |
|  | 400 | 0.0215  [-0.00111, 0.0441] | 0.258  [0.242, 0.274] | 0.263  [0.262, 0.265] | 95.4  [93.6, 97.2] | 4.92  [4.91, 4.92] |
|  | 10,000 | 0.0227  [0.0000335, 0.0454] | 0.259  [0.243, 0.275] | 0.264  [0.263, 0.265] | 95.6  [93.8, 97.4] | 150  [148, 151] |

**Supplementary table 3:** Scenario B: Continuous $Y$, continuous $X$, and continuous $U=\left( U_{1},U_{2} \right)$, summary of results for $\beta_{X}$ from the Monte Carlo quantitative bias analysis with differing number of replications, $m$: bias and empirical standard error of the point estimate, mean of model standard error, coverage of 95% interval estimate, and mean runtime. [95% Monte Carlo interval]

| **Prior** | $\boldsymbol{m}$ | **Bias** | **Empirical standard error** | **Mean**  **standard error** | **Coverage %** | **Mean runtime**  **in seconds** |
| --- | --- | --- | --- | --- | --- | --- |
| Very informative; Uniform prior for $\eta_{U_{1}}, \eta_{U_{2}}$ | 100 | -0.00311  [-0.00882, 0.00260] | 0.0652  [0.0611, 0.0692] | 0.0739  [0.0734, 0.0744] | 96.8  [95.3, 98.3] | 0.661  [0.658, 0.664] |
|  | 400 | -0.00338  [-0.00904, 0.00229] | 0.0646  [0.0606, 0.0686] | 0.0738  [0.0735, 0.0742] | 97.0  [95.5, 98.5] | 2.65  [2.63, 2.67] |
|  | 10,000 | -0.00349  [-0.00912, 0.00214] | 0.0642  [0.0603, 0.0682] | 0.0738  [0.0736, 0.0739] | 96.2  [94.5, 97.9] | 72.5  [71.9, 73.0] |
| Informative; Uniform prior for $\eta_{U_{1}}, \eta_{U_{2}}$ | 100 | -0.00227  [-0.00796, 0.00341] | 0.0649  [0.0608, 0.0689] | 0.0827  [0.0822, 0.0833] | 98.4  [97.3, 99.5] | 1.16  [1.15, 1.17] |
|  | 400 | -0.00271  [-0.00835, 0.00293] | 0.0643  [0.0603, 0.0683] | 0.0831  [0.0828, 0.0834] | 98.6  [97.6, 99.6] | 4.53  [4.49, 4.57] |
|  | 10,000 | -0.00283  [-0.00846, 0.00280] | 0.0642  [0.0602, 0.0682] | 0.0832  [0.0830, 0.0834] | 99.0  [98.1, 99.9] | 121  [120, 122] |
| Informative; Gamma prior for $1/{\eta_{U_{1}}^{2}}, 1/{\eta_{U_{2}}^{2}}$ | 100 | -0.00252  [-0.00823, 0.00319] | 0.0651  [0.0611, 0.0692] | 0.0833  [0.0827, 0.0838] | 97.8  [96.5, 99.1] | 0.745  [0.737, 0.753] |
|  | 400 | -0.00304  [-0.00872, 0 .00264] | 0.0648  [0.0608, 0.0688] | 0.0836  [0.0833, 0.0839] | 98.2  [97.0, 99.4] | 2.61  [2.59, 2.62] |
|  | 10,000 | -0.00275  [-0.00838, 0.00287] | 0.0642  [0.0602, 0.0681] | 0.0835  [0.0833, 0.0836] | 98.8  [97.8, 99.8] | 71.9  [70.9, 72.9] |

### **5. Choosing the numbers of burn-in and monitoring iterations for the Bayesian QBA**

For each scenario, we chose values for the numbers of burn-ins and monitoring iterations based on running standard convergence checks [6] using the informative prior setting. We generated a long chain of 50,000 iterations on one dataset and selected 1,000 burn-in and 3,000 monitoring iterations based on the trace plot of the Markov chain for the exposure effect $\beta_{X}$. The length of the burn-in period was chosen such that the effect of initial values on the point estimates for $\hat{\beta}_{X}$ is no longer present. The length of the monitoring period was chosen such that increasing it would not change the median, standard error and 2.5^th^ and 97.5^th^ percentiles of $\hat{\beta}_{X}$. For all parameters, initial values were chosen randomly by sampling from $N\left( {0, 1}^{2} \right)$ if they are coefficients, a uniform distribution within the allowable range for $\eta_{U_{1}}$ and $\eta_{U_{2}}$, and a $Gamma \left( 0.1, 1 \right)$ (i.e., shape of 0.1 and rate of 1) for the residual precision of linear regression $1/{\varepsilon_{Y}^{2}}$ when $Y$ is continuous. We verified our choices were not dependent on a particular set of initial values by generating three additional chains with different sets of initial values. We then verified the chosen burn-in and monitoring periods were sufficient for other datasets by generating three chains on nine other datasets and ensuring the Rhat from each dataset is less than 1.05 [7] and the effective sample size of each individual chain is less than our chosen number of monitoring iterations [6].

### **6. Results of simulation study I: Monte Carlo versus fully Bayesian implementation**

**Supplementary table 4:** Scenario A: Binary $Y$, binary $X$, and continuous $U=\left( U_{1} \right)$, summary of results for $\beta_{X}$ from the Monte Carlo and Bayesian quantitative bias analyses (QBAs): bias and empirical standard error of the point estimate, mean of model standard error, coverage of 95% interval estimate, and mean runtime. [95% Monte Carlo interval]

| **Method** | **Prior** | **Bias** | **Empirical standard error** | **Mean**  **standard error** | **Coverage %** | **Mean runtime**  **in seconds** |
| --- | --- | --- | --- | --- | --- | --- |
| Full analysis | - | 0.0139  [-0.00881, 0.0366] | 0.259  [0.243, 0.275] | 0.248  [0.246, 0.249] | 94.6  [92.6, 96.6] | - |
| Naïve analysis | - | 0.433  [0.412, 0.455] | 0.245  [0.230, 0.260] | 0.235  [0.233, 0.236] | 56.6  [52.3, 60.9] | - |
| Monte Carlo QBA^$^ | Very informative; Uniform prior for $\eta_{U_{1}}$ | 0.0175  [-0.00528, 0.0403] | 0.260  [0.244, 0.276] | 0.248  [0.246, 0.251] | 95.4  [93.6, 97.2] | 1.30  [1.29, 1.32] |
|  | Informative; Uniform prior for $\eta_{U_{1}}$ | 0.0218  [-0.000839, 0.0445] | 0.259  [0.243, 0.275] | 0.263  [0.260, 0.265] | 95.0  [93.1, 96.9] | 1.32  [1.31, 1.33] |
|  | Informative; Gamma prior for $1/{\eta_{U}^{2}}$ | 0.0237  [0.000761, 0.0466] | 0.262  [0.245, 0.278] | 0.264  [0.262, 0.267] | 95.6  [93.8, 97.4] | 1.25  [1.24, 1.25] |
| Bayesian QBA^$^ | Very informative; Uniform prior for $\eta_{U}$ | 0.0287  [0.00593, 0.0514] | 0.259  [0.243, 0.276] | 0.249  [0.248, 0.251] | 94.2  [92.2, 96.2] | 28.9  [28.5, 29.3] |
|  | Informative; Uniform prior for $\eta_{U}$ | 0.0266  [0.00389, 0.0493] | 0.259  [0.243, 0.275] | 0.266  [0.264, 0.267] | 95.2  [93.3, 97.1] | 29.0  [28.6, 29.4] |
|  | Informative; Gamma prior for $1/{\eta_{U}^{2}}$ | 0.0276  [0.00486, 0.0504] | 0.260  [0.244, 0.276] | 0.266  [0.264, 0.267] | 95.2  [93.3, 97.1] | 27.4  [27.1, 27.8] |

$ QBA using bias model of *qbaconfound* shown in eq:2 of the main paper.

**Supplementary table 5:** Scenario B: Continuous $Y$, continuous $X$, and continuous $U=\left( U_{1},U_{2} \right)$, summary of results for $\beta_{X}$ from the Monte Carlo and Bayesian quantitative bias analyses (QBAs): bias and empirical standard error of the point estimate, mean of model standard error, coverage of 95% interval estimate, and mean runtime. [95% Monte Carlo interval]

| **Method** | **Prior** | **Bias** | **Empirical standard error** | **Mean**  **standard error** | **Coverage %** | **Mean runtime**  **in seconds** |
| --- | --- | --- | --- | --- | --- | --- |
| Full analysis | - | -0.00145  [-0.00718, 0.00427] | 0.0653  [0.0613, 0.0694] | 0.0677  [0.0675, 0.0679] | 95.2  [93.3, 97.1] | - |
| Naïve analysis | - | 0.246  [0.241, 0.252] | 0.0642  [0.0602, 0.0682] | 0.0664  [0.0662, 0.0666] | 3.40  [1.81, 4.99] | - |
| Monte Carlo QBA^$^ | Very informative; Uniform prior for $\eta_{U_{1}}, \eta_{U_{2}}$ | -0.00311  [-0.00882, 0.00260] | 0.0652  [0.0611, 0.0692] | 0.0739  [0.0734, 0.0744] | 96.8  [95.3, 98.3] | 0.661  [0.658, 0.664] |
|  | Informative; Uniform prior for $\eta_{U_{1}}, \eta_{U_{2}}$ | -0.00227  [-0.00796, 0.00341] | 0.0649  [0.0608, 0.0689] | 0.0827  [0.0822, 0.0833] | 98.4  [97.3, 99.5] | 1.16  [1.15, 1.17] |
|  | Informative; Gamma prior for $1/{\eta_{U_{1}}^{2}}, 1/{\eta_{U_{2}}^{2}}$ | -0.00252  [-0.00823, 0.00319] | 0.0651  [0.0611, 0.0692] | 0.0833  [0.0827, 0.0838] | 97.8  [96.5, 99.1] | 0.745  [0.737, 0.753] |
| Bayesian  QBA^$^ | Very informative; Uniform prior for $\eta_{U_{1}}, \eta_{U_{2}}$ | -0.00381  [-0.00943, 0.00181] | 0.0641  [0.0601, 0.0681] | 0.0678  [0.0676, 0.0680] | 95.2  [93.3, 97.1] | 8.40  [8.34, 8.45] |
|  | Informative; Uniform prior for $\eta_{U_{1}}, \eta_{U_{2}}$ | -0.00407  [-0.00969, 0.00156] | 0.0641  [0.0601, 0.0681] | 0.0779  [0.0774, 0.0783] | 97.6  [96.3, 98.9] | 8.58  [8.52, 8.63] |
|  | Informative; Gamma prior for $1/{\eta_{U_{1}}^{2}}, 1/{\eta_{U_{2}}^{2}}$ | -0.00176  [-0.00906, 0.00553] | 0.0832  [0.0781, 0.0884] | 0.0809  [0.0760, 0.0859] | 95.8  [94.0, 97.6] | 5.38  [5.36, 5.41] |

$ QBA using bias model of *qbaconfound* shown in eq:2 of the main paper.

### **7. Results of simulation study II: Varying the associations between the confounders**

**Supplementary table 6:** Scenario A: Binary $Y$, binary $X$, and continuous $U=\left( U_{1} \right)$ with different levels of association between the confounders, summary of results for $\beta_{X}$: bias and empirical standard error of the point estimate, mean of its standard error, and coverage of its 95% interval estimate. Monte Carlo QBA applied with very informative priors and 100 replications. [95% Monte Carlo interval]

| **U-C associations** | **Method** | **Bias** | **Empirical standard error** | **Mean**  **standard error** | **Coverage %** |
| --- | --- | --- | --- | --- | --- |
| Independent $\left( \zeta=0 \right)$ | Full | 0.00478  [-0.0171, 0.0266] | 0.249  [0.234, 0.265] | 0.246  [0.245, 0.247] | 95.2  [93.3, 97.1] |
|  | Naïve | 0.429  [0.409, 0.449] | 0.233  [0.218, 0.247] | 0.233  [0.232, 0.234] | 56.8  [52.5, 61.1] |
|  | Monte Carlo | 0.0146  [-0.00711, 0.0364] | 0.248  [0.233, 0.264] | 0.246  [0.244, 0.248] | 96.0  [94.3, 97.7] |
| Observed $\left( \zeta=1 \right)$ | Full | 0.0139  [-0.00881, 0.0366] | 0.259  [0.243, 0.275] | 0.248  [0.246, 0.249] | 94.6  [92.6, 96.6] |
|  | Naïve | 0.433  [0.412, 0.455] | 0.245  [0.230, 0.260] | 0.235  [0.233, 0.236] | 56.6  [52.3, 60.9] |
|  | Monte Carlo | 0.0175  [-0.00528, 0.0403] | 0.260  [0.244, 0.276] | 0.248  [0.246, 0.251] | 95.4  [93.6, 97.2] |
| Strong $\left( \zeta=2 \right)$ | Full | 0.00707  [-0.0146, 0.0287] | 0.247  [0.232, 0.262] | 0.249  [0.248, 0.251] | 95.2  [93.3, 97.1] |
|  | Naïve | 0.428  [0.407, 0.448] | 0.235  [0.220, 0.249] | 0.236  [0.235, 0.238] | 55.0  [50.6, 59.4] |
|  | Monte Carlo | 0.0134  [-0.00837, 0.0352] | 0.248  [0.233, 0.264] | 0.251  [0.248, 0.253] | 95.4  [93.6, 97.2] |

**Supplementary table 7:** Scenario B: Continuous $Y$, continuous $X$, and continuous $U=\left( U_{1},U_{2} \right)$ with different levels of association between the confounders, summary of results for $\beta_{X}$: bias and empirical standard error of the point estimate, mean of its standard error, and coverage of its 95% interval estimate. Monte Carlo QBA applied with very informative priors and 100 replications. [95% Monte Carlo interval]

| $\boldsymbol{U-C}$ **and** $\boldsymbol{U}_{\boldsymbol{1}}\boldsymbol{-}\boldsymbol{U}_{\boldsymbol{2}}$ **associations** | **Method** | **Bias** | **Empirical standard error** | **Mean**  **standard error** | **Coverage %** |
| --- | --- | --- | --- | --- | --- |
| Independent $\left( \zeta=0 \right)$ | Full | 0.00366  [-0.00235, 0.00966] | 0.0685  [0.0643, 0.0728] | 0.0679  [0.0677, 0.0681] | 95.4  [93.6, 97.2] |
|  | Naïve | 0.251  [0.245, 0.257] | 0.0675  [0.0633, 0.0716] | 0.0663  [0.0661, 0.0665] | 2.80  [1.35, 4.25] |
|  | Monte Carlo | 0.00134  [-0.00468, 0.00735] | 0.0686  [0.0644, 0.0729] | 0.0737  [0.0732, 0.0742] | 97.2  [95.8, 98.6] |
| Standard $\left( \zeta=1 \right)$ | Full | -0.00145  [-0.00718, 0.00427] | 0.0653  [0.0613, 0.0694] | 0.0677  [0.0675, 0.0679] | 95.2  [93.3, 97.1] |
|  | Naïve | 0.246  [0.241, 0.252] | 0.0642  [0.0602, 0.0682] | 0.0664  [0.0662, 0.0666] | 3.40  [1.81, 4.99] |
|  | Monte Carlo | -0.00311  [-0.00882, 0.00260] | 0.0652  [0.0611, 0.0692] | 0.0739  [0.0734, 0.0744] | 96.8  [95.3, 98.3] |
| Strong $\left( \zeta=2 \right)$ | Full | -0.00101  [-0.00703, 0.00501] | 0.0687  [0.0644, 0.0729] | 0.0676  [0.0674, 0.0677] | 94.4  [92.4, 96.4] |
|  | Naïve | 0.250  [0.244, 0.256] | 0.0662  [0.0621, 0.0703] | 0.0666  [0.0664, 0.0667] | 3.80  [2.12, 5.48] |
|  | Monte Carlo | 0.000173  [-0.00569, 0.00603] | 0.0669  [0.0627, 0.0710] | 0.0737  [0.0732, 0.0742] | 97.2  [95.8, 98.6] |

### **8. Simulation study III: Binary unmeasured confounder(s)**

#### 8.1 Design of simulation study III

This simulation study follows a similar design to simulation study II. We considered two scenarios:

Scenario C: binary outcome $Y$, binary exposure $X$, and a single binary unmeasured confounder $U=\left( U_{1} \right)$, and
Scenario D: continuous outcome $Y$, continuous exposure $X$, and two binary unmeasured confounders $U=\left( U_{1},U_{2} \right)$.

In both scenarios, there are measured confounders $C=\left( {C_{con},C}_{bin},C_{cat} \right)$ as described in supplementary section 3.1 and we varied the magnitude of the associations between $U$ and $C$, and between $U_{1}$ and $U_{2}$ (scenario D only). We compared a full data analysis (where $U$ is treated as measured), a naïve analysis (that ignores $U$), and a Monte Carlo QBA using *qbaconfound* with very informative priors and 100 replications.

##### 8.1.1 Scenario C: binary outcome, binary exposure, single binary unmeasured confounder

Given data on measured confounders $C=\left( {C_{con},C}_{bin},C_{cat} \right)$ (simulated as described in supplementary section 3.1), data on $Y,X,$ and $U=\left( U_{1} \right)$ were generated as follows:

$Pr\left( X=1 \right)=expit\left\{ -4.819-0.3487C_{cat1}-0.7207 C_{cat2}+0.4609 C_{bin}+1.024 C_{con} \right\}$,

$Pr\left( U=1 \right)=expit\left\{ \alpha_{0\zeta}-\left( \zeta\times0.1639 \right)C_{cat1}-\left( \zeta\times0.0004130 \right)C_{cat2}+\left( \zeta\times0.04386 \right) C_{bin}+\left( \zeta\times0.6078 \right) C_{con}+1.178X \right\}$,

$Pr\left( Y=1 \right)=expit\left\{ -2.37+0.1753 C_{cat1}+0.04492C_{cat2}-0.3559C_{bin}-0.009095C_{con}+0 X+1.5 U \right\}$,

where $\zeta$ influences the magnitude of the associations between $U$ and $C$. We considered three values for $\zeta$ and intercept term $\alpha_{0\zeta}$ was chosen such that marginal prevalence of $U$ was approximately 20.5%:

- $\zeta=0$ encodes independence between $U$ and $C$ given $X$ and $\alpha_{0\zeta}=-1.65$.
- $\zeta=1$ encodes observed associations between $U$ and $C$ and $\alpha_{0\zeta}=-3.633$.
- $\zeta=2$ encodes strong associations between $U$ and $C$ and $\alpha_{0\zeta}=-5.65$.

##### 8.1.2 Scenario D: Continuous outcome, continuous exposure, two binary unmeasured confounders

Given data on measured confounders $C=\left( {C_{con},C}_{bin},C_{cat} \right)$ (simulated as described in supplementary section 3.1), data on $Y,X,$ and $U=(U_{1},U_{2})$ were generated as follows:

$X\sim N\left( 13.41-0.1988 C_{cat1}-0.3895 C_{cat2}+0.1442 C_{bin}+1.719 C_{con}, {2.176}^{2} \right)$,

$Pr\left( U_{1}=1 \right)=expit\left\{ \alpha_{1,0\zeta}-\left( \zeta\times0.1704 \right)C_{cat1}-\left( \zeta\times0.02826 \right)C_{cat2}+\left( \zeta\times0.1006 \right) C_{bin}+\left( \zeta\times0.4337 \right) C_{con}+0.2500 X \right\}$,

$Pr\left( U_{2}=1 \right)=expit\left\{ \alpha_{2,0\zeta}-\left( \zeta\times0.08843 \right)C_{cat1}-\left( \zeta\times0.1626 \right)C_{cat2}+\left( \zeta\times0.007311 \right) C_{bin}+\left( \zeta\times0.02073 \right) C_{con}+0.2500 X +\left( \zeta\times0.1495 \right)U_{1} \right\}$,

$Y\sim N\left( 8.534+0.5421C_{cat1}+0.5630C_{cat2}-0.2531C_{bin}-0.1783 C_{con}+0\times X+2.500 U_{1}+1.500 U_{2}, {4.322}^{2} \right)$,

where $\zeta$ influences the magnitude of the associations between $U$ and $C$, and between $U_{1}$ and $U_{2}$. We considered three values for $\zeta$ and the intercept terms $\alpha_{1,0\zeta}$ and $\alpha_{2,0\zeta}$ were chosen such that marginal prevalences of $U_{1}$ and $U_{2}$ were approximately 14.10% and 17.98%, respectively:

- $\zeta=0$ encodes independence between $U$ and $C$ given $X$, and between $U_{1}$ and $U_{2}$ given $X$ and $C$, with $\alpha_{1,0\zeta}=-6.695$ and $\gamma_{0\zeta}=-6.395$.
- $\zeta=1$ encodes observed associations between $U$ and $C$, and between $U_{1}$ and $U_{2}$ with $\alpha_{1,0\zeta}=-8.118$ and $\gamma_{0\zeta}=-6.406$.
- $\zeta=2$ encodes strong associations between $U$ and $C$, and between $U_{1}$ and $U_{2}$ with $\alpha_{1,0\zeta}=-9.563$ and $\gamma_{0\zeta}=-6.420$.

#### 8.2 Results of simulation study III

Supplementary tables 8 and 9 summarise the results of the simulation study for exposure effect $\beta_{X}$ when data were generated under scenario C (binary $Y$, $X$, and $U=\left( U_{1} \right)$), and scenario D (continuous $Y$ and $X$, and binary $U=\left( U_{1},U_{2} \right)$), respectively. As expected, the full data analysis resulted in unbiased estimates with close to nominal CI coverage whilst the naïve analysis resulted in biased estimates with substantial CI undercoverage. Applying *qbaconfound* with very informative priors (virtually) eliminated the bias due to unmeasured confounding and interval coverage was close the to nominal level. The performance of *qbaconfound* was not affected by the strength of the associations between $U$ and $C$ (and where relevant between $U_{1}$ and $U_{2}$).

**Supplementary table 8:** Scenario C: For binary $Y$, binary $X$, and binary $U=\left( U_{1} \right)$ with different levels of association between the confounders, summary of results for $\beta_{X}$: bias and empirical standard error of the point estimate, mean of its standard error, and coverage of its 95% interval estimate. Monte Carlo QBA applied with very informative priors and 100 replications. [95% Monte Carlo interval]

| $\boldsymbol{U-C}$ **associations** | **Method** | **Bias** | **Empirical standard error** | **Mean**  **standard error** | **Coverage %** |
| --- | --- | --- | --- | --- | --- |
| Independent $\left( \zeta=0 \right)$ | Full | 0.00603  [-0.0153, 0.0274] | 0.243  [0.228, 0.259] | 0.250  [0.248, 0.251] | 95.2  [93.3, 97.1] |
|  | Naïve | 0.396  [0.377, 0.416] | 0.223  [0.209, 0.237] | 0.233  [0.232, 0.234] | 60.2  [55.9, 64.5] |
|  | Monte Carlo | 0.0156  [-0.00570, 0.0369] | 0.243  [0.228, 0.258] | 0.253  [0.251, 0.255] | 96.0  [94.3, 97.7] |
| Observed $\left( \zeta=1 \right)$ | Full | -0.0126  [-0.0342, 0.00897] | 0.246  [0.231, 0.261] | 0.250  [0.249, 0.251] | 94.8  [92.9, 96.7] |
|  | Naïve | 0.379  [0.359, 0.400] | 0.233  [0.218, 0.247] | 0.233  [0.232, 0.234] | 62.0  [57.7, 66.3] |
|  | Monte Carlo | -0.00325  [-0.0253, 0.0188] | 0.252  [0.236, 0.267] | 0.253  [0.251, 0.255] | 95.0  [93.1, 96.9] |
| Strong $\left( \zeta=2 \right)$ | Full | -0.00471  [-0.0276, 0.0182] | 0.261  [0.245, 0.277] | 0.250  [0.248, 0.251] | 95.0  [93.1, 96.9] |
|  | Naïve | 0.372  [0.351, 0.392] | 0.234  [0.220, 0.249] | 0.233  [0.232, 0.234] | 60.2  [55.9, 64.5] |
|  | Monte Carlo | -0.00935  [-0.0315, 0.0128] | 0.252  [0.237, 0.268] | 0.255  [0.253, 0.257] | 96.0  [94.3, 97.7] |

**Supplementary table 9:** Scenario D: For continuous $Y$, continuous $X$, and binary $U=\left( U_{1},U_{2} \right)$ with different levels of association between the confounders, summary of results for $\beta_{X}$: bias and empirical standard error of the point estimate, mean of its standard error, and coverage of its 95% interval estimate. Monte Carlo QBA applied with very informative priors and 100 replications. [95% Monte Carlo interval].

| $\boldsymbol{U-C}$ **associations** | **Method** | **Bias** | **Empirical standard error** | **Mean**  **standard error** | **Coverage %** |
| --- | --- | --- | --- | --- | --- |
| Independent $\left( \zeta=0 \right)$ | Full | 0.00493  [-0.00127, 0.0111] | 0.0708  [0.0664, 0.0751] | 0.0653  [0.0651, 0.0654] | 94.0  [91.9, 96.1] |
|  | Naïve | 0.129  [0.123, 0.136] | 0.0705  [0.0661, 0.0749] | 0.0646  [0.0644, 0.0648] | 49.0  [44.6, 53.4] |
|  | Monte Carlo QBA | -0.00411  [-0.00214, 0.0104] | 0.0712  [0.0668, 0.0757] | 0.0687  [0.0683, 0.0692] | 94.2  [92.2, 96.2] |
| Observed $\left( \zeta=1 \right)$ | Full | -0.00176  [-0.00766, 0.00414] | 0.0673  [0.0632, 0.0715] | 0.0653  [0.0651, 0.0655] | 94.8  [92.9, 96.7] |
|  | Naïve | 0.123  [0.117, 0.129] | 0.0653  [0.0613, 0.0694] | 0.0647  [0.0645, 0.0648] | 50.2  [45.8, 54.6] |
|  | Monte Carlo QBA | -0.00254  [-0.00830, 0.00322] | 0.0657  [0.0617, 0.0698] | 0.0688  [0.0683, 0.0692] | 96.6  [95.0, 98.2] |
| Strong $\left( \zeta=2 \right)$ | Full | -0.00558  [-0.0113, 0.000122] | 0.0650  [0.0610, 0.0691] | 0.0654  [0.0652, 0.0656] | 94.8  [92.9, 96.7] |
|  | Naïve | 0.120  [0.115, 0.126] | 0.0648  [0.0608, 0.0689] | 0.0649  [0.0647, 0.0650] | 55.2  [50.8, 59.6] |
|  | Monte Carlo QBA | -0.00583  [-0.0116, -0.000108] | 0.0653  [0.0613, 0.0694] | 0.0687  [0.0682, 0.0692] | 96.2  [94.5, 97.9] |

### **9. Results of applying *qbaconfound* in other scenarios**

#### 9.1 Design of simulation study IV

We further evaluated *qbaconfound* when the substantive analysis was a:

Scenario E: linear regression with a categorical exposure,
Scenario F: multinomial logistic regression with continuous exposure, and
Scenario G: Cox proportional hazard regression with a continuous exposure.

Note that in scenario E, $U$ consisted of two unmeasured confounders, one continuous and one binary, and in scenarios F and G, $U$ was a single continuous unmeasured confounder.

For all scenarios, we generated 500 simulated datasets, each with 1,000 observations. We compared a full data analysis (where $U$ was treated as measured), a naïve analysis (that ignores $U$), and a Monte Carlo QBA using *qbaconfound* with very informative priors (as defined in the main paper) and 100 Monte Carlo replications. Below we describe the data generating models for $Y,X,$ and $U$. With the exception of scenario E, measured confounders $C=\left( {C_{con},C}_{bin},C_{cat} \right)$ were simulated as described in supplementary section 3.1.

##### 9.1.1 Scenario E: Continuous outcome, categorical exposure, continuous and binary unmeasured confounders

Consider the scenario in which $Y$ is continuous, $X$ is a 3-level categorical exposure, the substantive analysis is a linear regression, and there are two unmeasured confounders, $U_{1}$ is continuous and $U_{2}$ is binary. Given data on measured confounders $C=\left( {C_{con},C}_{bin},C_{cat} \right)$, data on $Y,X$ (baseline category 2), and $U=(U_{1},U_{2})$ were generated as follows:

$\frac{Pr\left( X=0 \right)}{Pr\left( X=2 \right)}=expit\left\{ 4.020+0.2612C_{cat1}+0.5005C_{cat2}-0.1836C_{bin}-2.173 C_{con} \right\}$,

$\frac{Pr\left( X=1 \right)}{Pr\left( X=2 \right)}=expit\left\{ 3.040+0.1508C_{cat1}+0.2860C_{cat2}-0.1018C_{bin}-1.205 C_{con} \right\}$,

$U_{1}\sim N\left( 60.67-1.039 C_{cat1}-0.3788 C_{cat2}+0.5880 C_{bin}+3.055 C_{con}-7.492 X_{0}-4.175X_{1}, {9.554}^{2} \right)$,

$$Pr\left( U_{2}=1 \right)=expit\left\{ -0.5634-0.08549C_{cat1}-0.1699 C_{cat2}+0.01151 C_{bin}+0.1163 C_{con}-1.084 X_{0}-0.5996 X_{1} +0.01258 U_{1} \right\}$$

$Y\sim N\left( 8.534+0.5421C_{cat1}+0.5630C_{cat2}-0.2531C_{bin}-0.1783 C_{con}+0\times X_{0}+0\times X_{1}+0.1299 U_{1}+0.5 U_{2}, {4.322}^{2} \right)$,

where $X_{0}$ and $X_{1}$ are dummy variables corresponding to $X=0$ and $X=1$, respectively.

##### 9.1.2 Scenario F: Nominal outcome, continuous exposure, and single continuous unmeasured confounder

Consider the scenario in which $Y$ is a 3-category nominal variable, $X$ is a continuous exposure, the substantive analysis is a multinomial logistic regression, and there is one continuous unmeasured confounder $U$. Given data on measured confounders $C=\left( {C_{con},C}_{bin},C_{cat} \right)$, data on $Y$ (baseline category 2), $X$, and $U=(U_{1})$ were generated as follows:

$X\left| C \right.\sim N\left( 13.41-0.1988 C_{cat1}-0.3895 C_{cat2}+0.1442 C_{bin}+1.719 C_{con}, {2.176}^{2} \right)$,

$U\left| X,C \right.\sim N\left( 58.11-0.4942 C_{cat1}-0.8886 C_{cat2}+0.05031 C_{bin}+0.1617 C_{con}+1.281 X, {10.30}^{2} \right)$,

$\frac{Pr\left( Y=0 | X,C,U \right)}{Pr\left( Y=2 | X,C,U \right)}=exp\left\{ -7+0\times X-0.1022 C_{cat1}-0.1217 C_{cat2}+0.04580C_{bin}-0.02349 C_{con}+0.1000 U \right\}$,

$\frac{Pr\left( Y=1 | X,C,U \right)}{Pr\left( Y=2 | X,C,U \right)}=exp\left\{ -7+0\times X-0.1959 C_{cat1}-0.2413 C_{cat2}+0.08650 C_{bin}-0.05253 C_{con}+0.1000 U \right\}$.

##### 9.1.3 Scenario G: Time-to-event outcome, continuous exposure, and single continuous unmeasured confounder

Consider the scenario in which $Y=\left( S,D \right)$ for survival time $S$ and event indicator $D$ (1 for events, 0 for censored observations), $X$ is a continuous exposure, the substantive analysis is a Cox proportional hazards regression, and there is one continuous unmeasured confounder $U$. The measured confounders consist of binary $C_{bin}$, continuous $C_{con}$, and 4-category variable $C_{cat}$ with dummy variables $C_{cat1}$, $C_{cat2}$, and $C_{cat3}$. Data were generated according to the following model with the parameters based on a trial of Azathioprine for primary biliary cholangitis [8]:

$C_{cat} \sim multinomial\left( p_{0}=0.25, p_{1}=0.25, p_{2}=0.25,p_{3}=0.25 \right)$,

where $p_{j}=Pr\left( C_{cat}=j \right)$ for $j=0,1,2,3$, and

$Pr\left( C_{bin}=1 \right)=0.2$,

$C_{con}\sim N\left( -0.058,{5.9}^{2} \right)$,

$X\left| C\sim N\left( -0.13+0.063C_{cat1}+0.084C_{cat2}+0.058C_{cat3}+0.50C_{bin}-0.014C_{con},{0.40}^{2} \right) \right.$,

$U\left| X,C \right.\sim N\left( 55-1.4 C_{cat1}-0.29 C_{cat2}-0.22C_{cat3}+ 0.85C_{bin}-0.54C_{con}-7.8X, {10}^{2} \right)$,

$$h\left( s|X,C,U \right)= h_{0}\left( s \right) exp\left( 0\times X_{con}-0.043 C_{con}+0.70 C_{bin}-0.27 C_{cat1}-0.13 C_{cat2}-0.019 C_{cat3}+0.04 U_{con} \right)$$

Note that the event time was drawn from the above hazard function and censoring time was drawn from $Uniform \left( 0, 12 \right)$. Of the outcome $Y=\left( S,D \right)$, $S$ was the minimum of the event time and censoring time, and $D$ indicated whether $S$ was derived from an event time $\left( D=1 \right)$ or a censoring time $\left( D=0 \right)$.

#### 9.2 Results of simulation study IV

**Supplementary table 10:** Scenario E: For continuous $Y$, 3-level categorical $X$, and continuous and binary $U=\left( U_{1},U_{2} \right)$, summary of results for $\beta_{X_{0}}$ and $\beta_{X_{1}}$: bias and empirical standard error of the point estimate, mean of model standard error, coverage of 95% interval estimate, and mean runtime. Monte Carlo quantitative bias analysis (QBA) applied with very informative priors and 100 replications. [95% Monte Carlo interval]

| **Method** | **Exposure coefficient** | **Bias** | **Empirical standard error** | **Mean**  **standard error** | **Coverage %** |
| --- | --- | --- | --- | --- | --- |
| Full model analysis | $\beta_{X_{0}}$ | -0.0179  [-0.0815, 0.0458] | 0.726  [0.681, 0.771] | 0.744  [0.739, 0.749] | 95.2  [93.3, 97.1] |
| Naïve analysis | $\beta_{X_{0}}$ | -1.14  [-1.21, -1.07] | 0.762  [0.714, 0.809] | 0.762  [0.757, 0.768] | 67.6  [63.5, 71.7] |
| Monte Carlo QBA^$^ | $\beta_{X_{0}}$ | -0.0262  [-0.0935, 0.0411] | 0.768  [0.720, 0.815] | 0.822  [0.814, 0.829] | 97.2  [95.8, 98.6] |
| Full model analysis | $\beta_{X_{1}}$ | -0.00273  [-0.0306, 0.0252] | 0.318  [0.298, 0.338] | 0.325  [0.324, 0.326] | 94.2  [92.2, 96.2] |
| Naïve analysis | $\beta_{X_{1}}$ | -0.625  [-0.653, -0.596] | 0.326  [0.306, 0.347] | 0.330  [0.329, 0.331] | 53.0  [48.6, 57.4] |
| Monte Carlo QBA^$^ | $\beta_{X_{1}}$ | -0.00265  [-0.0314, 0.0261] | 0.328  [0.308, 0.348] | 0.356  [0.353, 0.358] | 96.6  [95.0, 98.2] |

$ QBA using bias model of *qbaconfound* shown in eq:2 of the main paper.

**Supplementary table 11:** Scenario F: For nominal $Y$, continuous $X$, and continuous $U=\left( U_{1} \right)$, summary of results for $\beta_{X}$: bias and empirical standard error of the point estimate, mean of model standard error, coverage of 95% interval estimate, and mean runtime. Monte Carlo quantitative bias analysis (QBA) applied with very informative priors and 100 replications. [95% Monte Carlo interval]

| **Method** | **Category of** $\boldsymbol{Y}$ | **Bias** | **Empirical standard error** | **Mean**  **standard error** | **Coverage %** |
| --- | --- | --- | --- | --- | --- |
| Full model analysis | 0 | -0.00630  [-0.0102, -0.00237] | 0.0449  [0.0421, 0.0477] | 0.0446  [0.0445, 0.0448] | 94.0  [91.9, 96.1] |
| Naïve analysis | 0 | 0.107  [0.103, 0.110] | 0.0399  [0.0374, 0.0424] | 0.0405  [0.0404, 0.0406] | 23.6  [19.9, 27.3] |
| Monte Carlo QBA^$^ | 0 | -0.00435  [-0.00834, -0.000353] | 0.0456  [0.0427, 0.0484] | 0.0474  [0.0471, 0.0478] | 96.0  [94.3, 97.7] |
| Full model analysis | 1 | -0.00420  [-0.00819, -0.000215] | 0.0455  [0.0426, 0.0483] | 0.0457  [0.0455, 0.0458] | 94.6  [92.6, 96.6] |
| Naïve analysis | 1 | 0.109  [0.105, 0.112] | 0.0411  [0.0385, 0.0436] | 0.0416  [0.0415, 0.0417] | 24.2  [20.4, 28.0] |
| Monte Carlo QBA^$^ | 1 | -0.00244  [-0.00658, 0.00170] | 0.0472  [0.0443, 0.0502] | 0.0479  [0.0476, 0.0482] | 96.0  [94.3, 97.7] |
|  | 2 | (Base category of $Y$) | | | |

$ QBA using bias model of *qbaconfound* shown in eq:2 of the main paper.

**Supplementary table 12:** Scenario G: Survival outcome $Y=\left( S,D \right)$, continuous $X$, and continuous $U=\left( U_{1} \right)$, summary of results for $\beta_{X}$: bias and empirical standard error of the point estimate, mean of model standard error, coverage of 95% interval estimate, and mean runtime. Monte Carlo quantitative bias analysis applied with very informative priors and 100 replications. [95% Monte Carlo interval]

| **Method** | **Bias** | **Empirical standard error** | **Mean**  **standard error** | **Coverage %** | **Mean runtime**  **in seconds** |
| --- | --- | --- | --- | --- | --- |
| Full model analysis | 0.00143  [-0.00955, 0.0124] | 0.125  [0.117, 0.133] | 0.122  [0.122, 0.123] | 94.4  [92.4, 96.4] | - |
| Naïve analysis | -0.291  [-0.302, -0.281] | 0.123  [0.115, 0.130] | 0.117  [0.116, 0.117] | 30.0  [26.0, 34.0] | - |
| Monte Carlo QBA^$^ | 0.0190  [0.00803, 0.0300] | 0.125  [0.117, 0.133] | 0.127  [0.126, 0.128] | 96.4  [94.8, 98.0] | 1.51  [1.50, 1.52] |

$ QBA using bias model of *qbaconfound* shown in eq:2 of the main paper.

### **10. Applied example**

**Supplementary table 13:** Prior distributions for the bias parameters of the applied example

| **Bias parameter** | **Prior distribution** |
| --- | --- |
| $\beta_{U}=\left( \beta_{U_{1}},\beta_{U_{2}} \right)$ | ${p(\beta}_{U_{1}})\sim N\left( 0.006931,{0.0002805}^{2} \right)$  $p(\beta_{U_{2}})\sim N\left( -1.312,{0.3453}^{2} \right)$ |
| $\alpha_{1,X}=\left( \alpha_{1,X_{never}},\alpha_{1,X_{former}}, \alpha_{1,X_{3to4}},\alpha_{1,X_{atleast5}} \right)$ | $p(\alpha_{1,X_{never}})\sim N\left( -43.47,{27.21}^{2} \right)$  $p(\alpha_{1,X_{former}})\sim N\left( 69.72,{18.63}^{2} \right)$  $p\left( \alpha_{1,X_{3to4}} \right)\sim N\left( -65.37,{18.07}^{2} \right)$  $p(\alpha_{1,X_{atleast5}})\sim N\left( -94.71,{19.64}^{2} \right)$ |
| $\eta_{U_{1}}$ | ${p(\eta}_{U_{1}})\sim Uniform\left( 372.6, 716.5 \right)$ |
| $\alpha_{2,X}=\left( \alpha_{2,X_{never}},{\alpha_{2,X_{former}},\alpha}_{2,X_{3to4}},\alpha_{2,X_{atleast5}} \right)$ | $p(\alpha_{2,X_{never}})\sim N\left( 0.4524,{0.1081}^{2} \right)$  $p(\alpha_{2,X_{former}})\sim N\left( -0.07963,{0.08107}^{2} \right)$  $p(\alpha_{2,X_{3to4}})\sim N\left( 0.3831,{0.07405}^{2} \right)$  ${p(\alpha}_{2,X_{atleast5}})\sim N\left( 0.3785,{0.07958}^{2} \right)$ |
| $\pi_{2}$ | $p\left( \pi_{2} \right)\sim Uniform\left( 0.2900, 0.3484 \right)$ |
| $\tilde{\alpha}_{2,0}$ | $p(\tilde{\alpha}_{2,0}) \sim N\left( -0.9175,{0.04286}^{2} \right)$ |

For all parameters in the Bayesian QBA, initial values were chosen randomly by sampling from $N\left( {0, 1}^{2} \right)$ for coefficients, or a uniform distribution within the allowable range for the residual standard deviation $\eta_{U_{1}},$ and a $Gamma \left( 0.1, 1 \right)$ for the residual precision of linear regression $1/{\varepsilon_{Y}^{2}}$.

**Supplementary table 14:** Point estimate with corresponding 95% interval estimate of the exposure effect $\beta_{X}$ for the mean difference in waist circumference (cm) compared to those who drank 1 to 2 drinks per drinking day

| **Method** | $\boldsymbol{\beta}_{\boldsymbol{X}_{\boldsymbol{never}}}$ | $\boldsymbol{\beta}_{\boldsymbol{X}_{\boldsymbol{former}}}$ | $\boldsymbol{\beta}_{\boldsymbol{X}_{\mathbf{3}\boldsymbol{to}\mathbf{4}}}$ | $\boldsymbol{\beta}_{\boldsymbol{X}_{\boldsymbol{atleast}\mathbf{5}}}$ |
| --- | --- | --- | --- | --- |
| Full analysis | 0.494  [-0.798, 1.79] | 1.53  [0.646, 2.42] | 0.655  [-0.204, 1.51] | 3.38  [2.44, 4.31] |
| Naïve analysis | 0.0697  [-1.27, 1.41] | 2.04  [1.12, 2.96] | 0.103  [-0.790, 0.995] | 2.62  [1.65, 3.59] |
| Monte Carlo QBA^$^ using prior $p\left( \pi_{2} \right)$ | 0.506  [-1.01, 2.01] | 1.54  [0.534, 2.55] | 0.668  [-0.322, 1.67] | 3.39  [2.35, 4.47] |
| Monte Carlo QBA^$^ using prior $p\left( \tilde{\alpha}_{2,0} \right)$ | 0.524  [-0.974, 2.01] | 1.53  [0.486, 2.55] | 0.660  [-0.326, 1.66] | 3.38  [2.31, 4.48] |
| Bayesian QBA^$^ using prior $p\left( \tilde{\alpha}_{2,0} \right)$ | 0.552  [-0.819, 1.96] | 1.60  [0.666, 2.54] | 0.735  [-0.205, 1.68] | 3.47  [2.46, 4.51] |

$ QBA using bias model of *qbaconfound* shown in eq:2 of the main paper.

**References**

| [1] | R. Hebdon, J. Stamey, D. Kahle and X. Zhang, “unmconf: an R package for Bayesian regression with unmeasured confounders,” *BMC Medical Research Methodology,* vol. 24, no. 195, pp. 1-10, 2024. |
| --- | --- |
| [2] | A. R. Willan and D. G. Watts, “Meaningful multicollinearity measures,” *Technometrics,* vol. 20, no. 4, pp. 407-412, 1978. |
| [3] | I. R. White, P. Royston and A. M. Wood, “Multiple imputation using chained equations: Issues and guidance for practice,” *Statistics in Medicine,* vol. 30, pp. 377-399, 2011. |
| [4] | T. P. Morris, I. R. White and M. J. Crowther, “Using simulation studies to evaluate statistical methods,” *Statistics in Medicine,* vol. 38, no. 11, pp. 2074-2102, 2019. |
| [5] | R. H. Keogh and T. P. Morris, “Multiple imputation in Cox regression when there are time-varying effects of covariates,” *Statistics in Medicine,* vol. 37, pp. 3661-3678, 2018. |
| [6] | A. Gelman, J. B. Carlin, H. S. Stern and D. B. Rubin, Bayesian data analysis, New York: Chapman and Hall/CRC, 2013. |
| [7] | A. Gelman and D. Rubin, “Inference from iterative simulation using multiple sequences,” *Statistical Science,* vol. 7, pp. 457-5111, 1992. |
| [8] | E. Christensen, J. Neuberger, J. Crowe, D. Altman, H. Popper, P. B, D. Doniach, L. Ranek, N. Tygstrup and R. Williams, “Beneficial effect of azathioprine and prediction of prognosis in primary biliary cirrhosis: Final results of an international trial,” *Gastroenterology,* vol. 89, pp. 1084-1091, 1985. |
